# Deciphering T-wave Morphologies on ECGs: The Simplified Egg and Changing Yolk Model and the Importance of the QTp Interval

**DOI:** 10.1101/2024.12.17.24318926

**Authors:** Kieran Stone, Arvind Mistry, Daniel Cyrus, John Cannon, Ignacy Mokrzecki, Faisal I Rezwan

## Abstract

The Electrocardiogram (ECG) serves as an integral tool in the diagnosis and management of a variety of cardiac diseases. It visualises electrical activity in the heart, offering insights into several cardiac processes, including ventricular repolarisation. The morphology of the T-wave observed on ECGs during this repolarisation phase varies and can be peaked, flat, inverted, or biphasic, each representing different cardiac conditions. Despite their prevalence, the interpretation of these patterns remains challenging. Therefore, we proposed the Simplified Egg and Changing Yolk Model, a novel idea to aid in the understanding of these T-wave morphologies in ECGs.

The proposed Simplified Egg and Changing Yolk Model was developed through an analysis of various T-wave morphologies and their corresponding clinical implications. The model was further designed to conceptualise the ST interval and the T-wave as a single unit, contributing to a simplified yet comprehensive understanding of ventricular repolarisation. In this context, the ‘Q-wave start to T-wave peak interval’(QTp) was compared to the more commonly used ‘corrected QT-interval’ (QTc) for assessing the risk of arrhythmia and the effects of medication that prolong the QT-interval.

The Simplified Egg and Changing Yolk Model could effectively explain and interpret the variation of ECG patterns associated with ventricular repolarisation. It provided insight into the relevance of deflections seen during this phase. Importantly, the model identified QTp as a more reliable measure than QTc for assessing arrhythmia risk and evaluating medication impacts on the QT-interval.

Our model offers a significant enhancement to the understanding of ventricular repolarisation and its manifestation on ECGs. By emphasising the superiority of QTp over QTc in clinical assessment, this model can have significant impact in clinical practice.

## Introduction

Electrocardiogram (ECG) signals provide significant information about a patient’s heart processes and are widely used to investigate both normal and pathological conditions of the heart^1^. These signals display the time evolution of the heart’s electrical activity for each heartbeat in the cardiac cycle, typically consisting of distinct depolarisation and repolarisation patterns^2^. These electrical activities in each cardiac cycle are detected by an ECG device and translates them into a graphical representation to identify anomalies in heart rhythm or any changes in heart morphology^3,4^. Any deviations in ECG traces from the ‘normal pattern’ (shown in Figure 1) are empirically interpreted by clinicians as ‘ECG abnormalities’.

**Figure 1:**
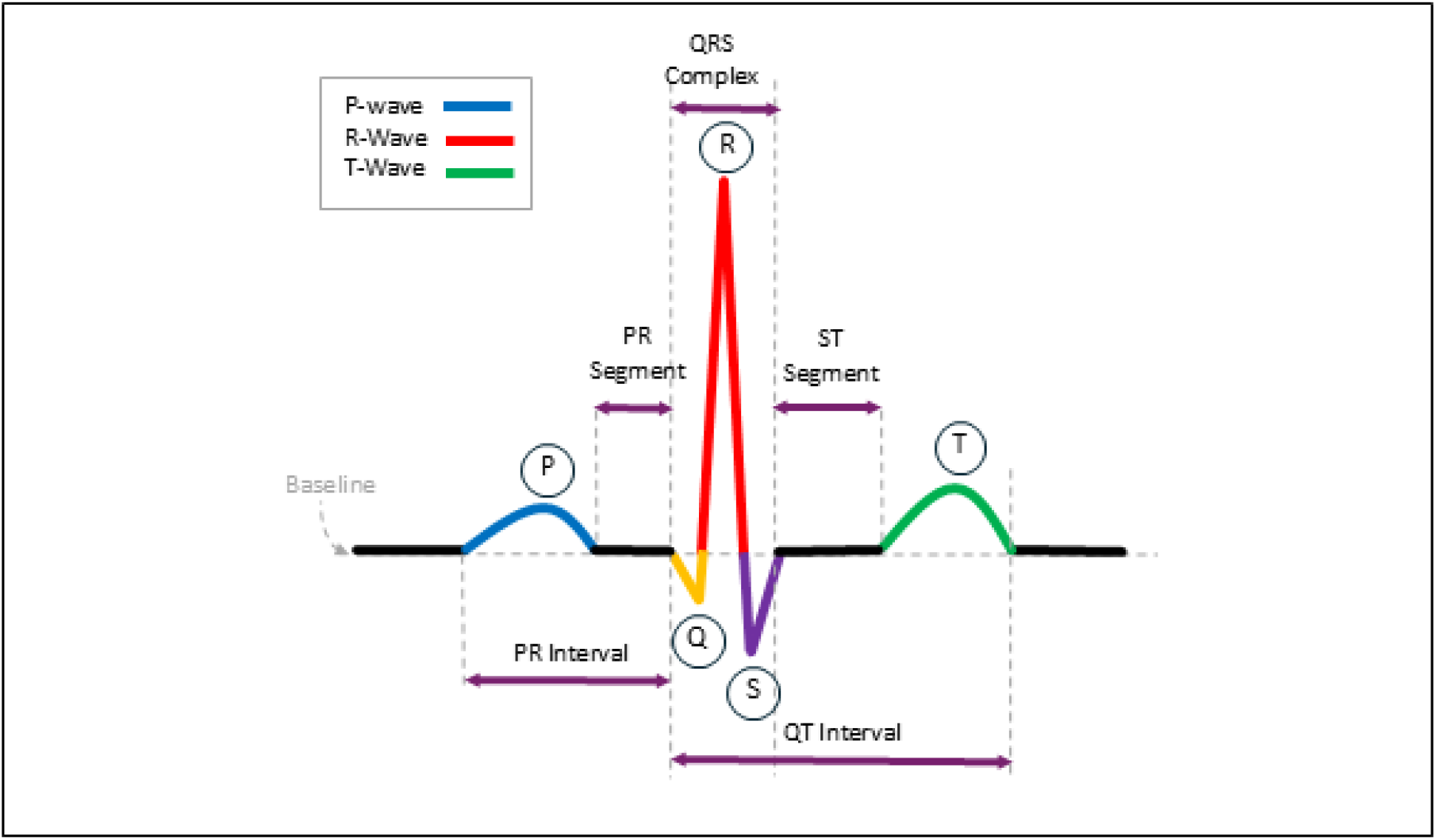
The classic ECG wave. The classic ECG wave illustrates the essential components of cardiac electrical activity, including the P-wave, QRS complex, and T-wave, as well as their respective intervals.

This knowledge has evolved over many years, with clinicians matching ECG patterns to pathological changes in the heart through experience. While this approach has been effective, it often lacks sufficient explanations for why certain patterns appear as they do, leading to the propagation of concepts without fully questioning the underlying reasons for the formation of these ECG traces.

Ventricular repolarisation as reflected in various T-wave morphologies on ECG traces, is a key aspect clinicians use to assess heart conditions. For instance, changes in the ST segment are crucial indicators of ischemia or infarct^5^. Despite their diagnostic importance, the underlying mechanisms causing ST elevation and ST depression remain unclear, with no definitive explanation provided for these phenomena^6^. Additionally, the absence of a definitive explanation for the typical ‘hump’ shape of the T-wave on an ECG, coupled with the unclear mechanisms behind various atypical T-wave morphologies such as peaked, inverted, flattened, or biphasic patterns, are particularly noteworthy^7,8^.

Beyond the limited understanding of the mechanisms behind various atypical T-wave morphologies, the QT interval is a crucial measurement on ECGs. Essentially, the QT interval reflects the time required for the heart’s ventricles to contract and then relax in preparation for the next heartbeat. This is crucial in evaluating heart function, as an abnormal duration may indicate various cardiac conditions, including arrhythmias^9^. A prolonged QT interval is associated with an increased risk of developing a potentially life-threatening arrhythmia known as Torsades de Pointes, while a shortened QT interval may also be linked to specific cardiac disorders^10,11^.

It is also noteworthy that the QT interval is influenced by heart rate^12^. Consequently, it is frequently corrected for the patient’s heart rate using different formulae (e.g. Bazett’s or Fridericia’s formulae), resulting in the corrected QT interval (QTc)^13^. However, several issues are associated with the current QTc interval when comparing cases of heart rate variability (for more details see Supplementary Material section 1).

This study explores ECG electrophysiology by focusing on its link to Wilson’s Central Terminal (WCT) and the origins of ECG deflections. We present a novel approach, the “Simplified Egg and Changing Yolk Model”, to clarify the ventricular repolarisation process seen in the T-wave. This model explains common T-wave abnormalities, enhancing the accuracy and effectiveness of QT interval analysis.

### The Simplified Egg and Changing Yolk Model

The “Simplified Egg and Changing Yolk Model” views the ventricular repolarisation process as occurring within a mass of heart muscle, simplified into an ‘egg’ shape. This shape influences the electrical deflection detected by ECG leads, depending on electrode position. In this example the green line represents the vector of lead V5, passing through the egg’s long axis. Signals detected to the right of the WCT for lead V5 register as positive, while those on the left register as negative. The resulting ECG trace is the net outcome of these positive and negative signals over time (see Figure 2)^14^.

**Figure 2:**
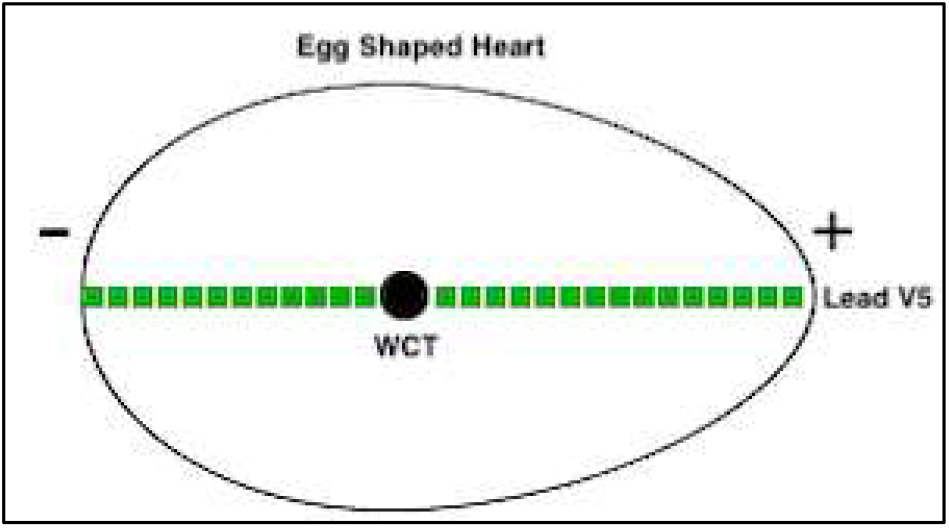
Representation of the heart in the Simplified Egg and Changing Yolk Model. This figure depicts the heart as an egg-shaped 3D object, with the WCT at its centre.

### Normal repolarisation sequence as explained by the model

In the Simplified Egg and Changing Yolk Model, the repolarisation signal expands from the WCT to the edges until the entire muscle mass has completed repolarisation. The expanding ‘yolk’ can be likened to ripples emanating from a stone dropped into still water, spreading out from the centre (WCT) in all directions but extending further on one side due to the asymmetrical egg shape. The model states that an ECG lead detects the one-dimensional repolarisation signal based on its vector through the egg shape. Signals moving in front of the WCT point produce a positive deflection, while those moving away cause a negative deflection^15^. The net result of these signals is registered on the ECG trace over time for that lead.

The sequence of the repolarising signal expanding out from the WCT to the outer boundaries of the egg shape is shown in Figure 3. The lead V5 vector is shown as a green line pointing into the egg through the WCT and in this example is looking from the right side of the egg shape. As the signal expands (shown in orange in the diagram) the net result from the positive and negative sides of the lead register as deflections (positive or negative) depending on which side has the greater signal amplitude. For a specified lead, this is represented as the following formula:

**Figure 3:**
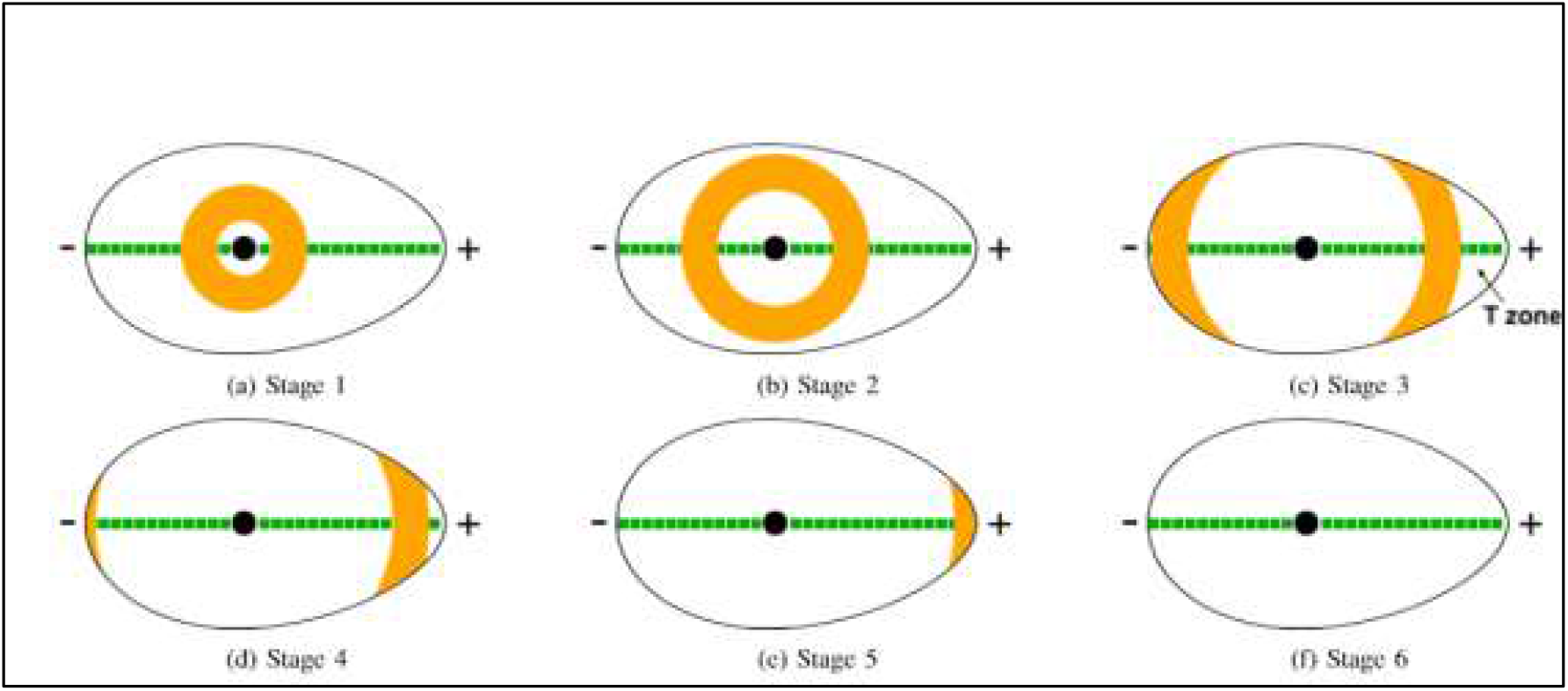
The repolarisation sequence of the heart. These diagrams show the progression of an expanding repolarisation wave from the WCT within the egg-shaped heart. The ‘T-zone’ refers to the asymmetric region of the egg shape, crucial in generating the T-wave in this lead.

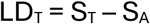

Here, LD_T_ is the deflection for a specified lead at time T, S_T_ represents the signal components moving towards the lead and S_A_ represents the signal components moving away from the lead at a given time.

The ‘components’ in this model consist of the signal itself, any enhancement of the signal caused by hypertrophy, and any damage in the muscle mass (location and severity).

As demonstrated in Figure 3, initially the positive and negative deflections in that lead balance out yielding a net isoelectric reading until the asymmetrical part of the egg shape is reached. This is the ST segment (Figures 3a to 3c). When the signal reaches the asymmetrical part of the egg shape, termed as the ‘T-zone’, an excess signal on one side which causes a deflection in the lead trace. In Figure 3d, this results in a rising positive deflection, which peaks when the signal’s front edge reaches the outer boundary of the ‘T-zone’ (Figure 3e). Eventually the repolarisation signal works its way out of the ‘T-zone’ of the egg shape (asymmetrical part) causing the positive deflection in the lead to return to the isoelectric line (Figure 3f). Once the signal has exited, the entire shape has become repolarised, ready for the next signal.

From the repolarisation sequence (shown in Figure 3) and the corresponding net deflections in Figure 4, the ECG trace stays on the isoelectric line (balanced) during the ST interval. A net positive signal is detected on the lead only when the repolarisation signal enters the asymmetrical portion of the egg shape (T-zone) and continues until the signal exits this boundary. In the proposed model, this ‘T-zone’ is responsible for generating the T-wave observed in the ECG. The time from the onset of repolarisation to the stage when the signal reaches the boundary of the ‘T-zone’ (Figures 3a to 3e) corresponds to the Q-wave start to T-wave peak (QTp) interval. In contrast, the commonly used corrected QT interval (QTc) spans from the start of the Q-wave to the point when the repolarisation signal has propagated out of the ‘T-zone’ (from 3a to 3f in the sequence). The QTp interval is marked when the repolarisation signal reaches the ‘T-zone’ boundary, while the QTc interval indicates when the signal exits the boundary. Comparing the QTc and QTp intervals, the QTp is more clearly defined and more reliably calculable.

**Figure 4:**
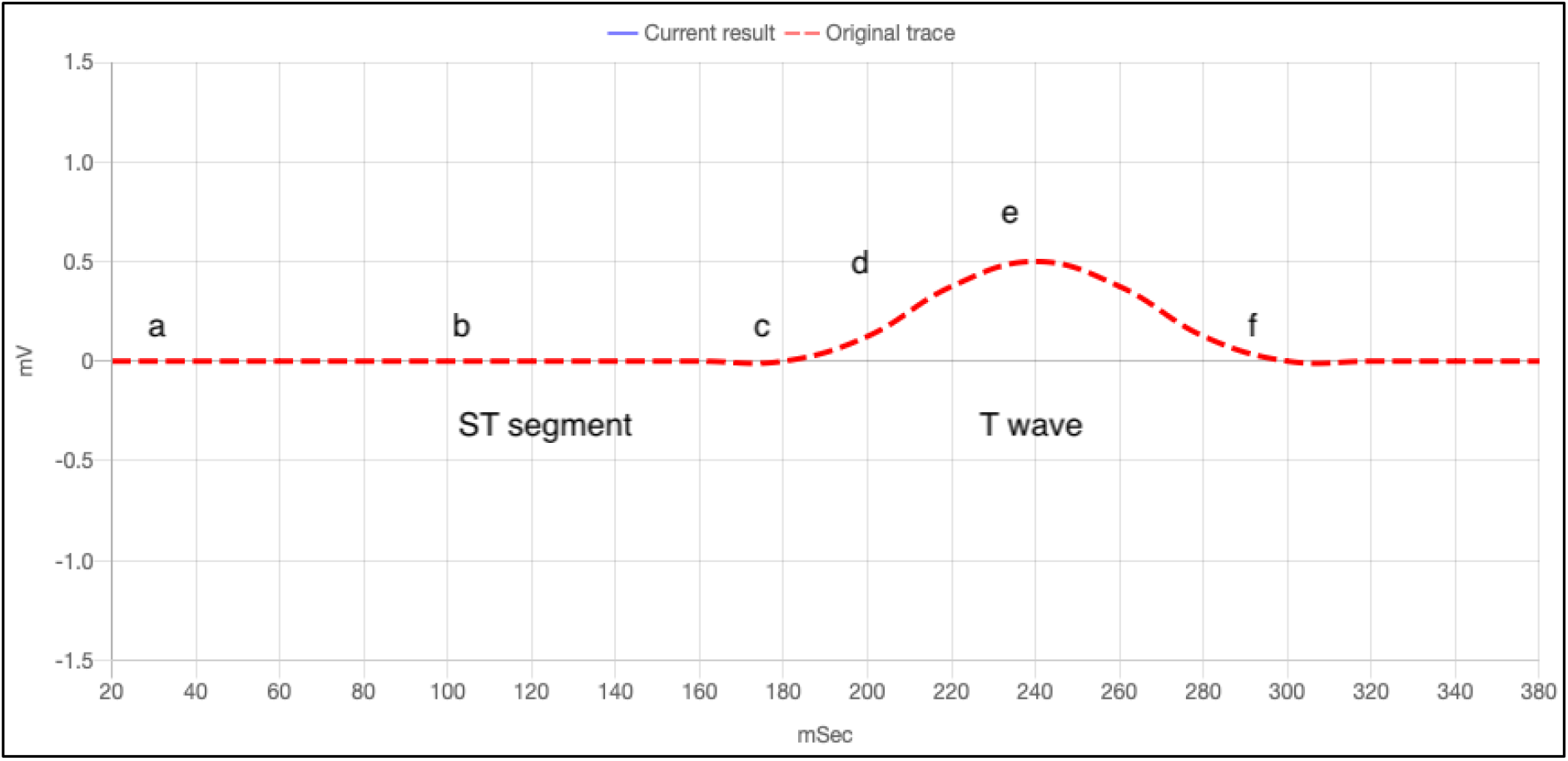
The ECG trace produced by repolarisation. This graph displays the ECG trace generated by the expanding repolarisation signal in the Simplified Egg and Changing Yolk Model. The letters correspond to the signal’s position in Figures 3a to 3f.

### Model Repolarisation Patterns with Heart Damaged Areas

The Simplified Egg and Changing Yolk Model offers a framework for studying the effects of damage to specific regions of heart that no longer contributes a repolarisation signal through the electrically inert area. In Figures 5a and 5c, the red areas represent where along the lead the damage, or non-electrical regions, are registered.

**Figure 5:**
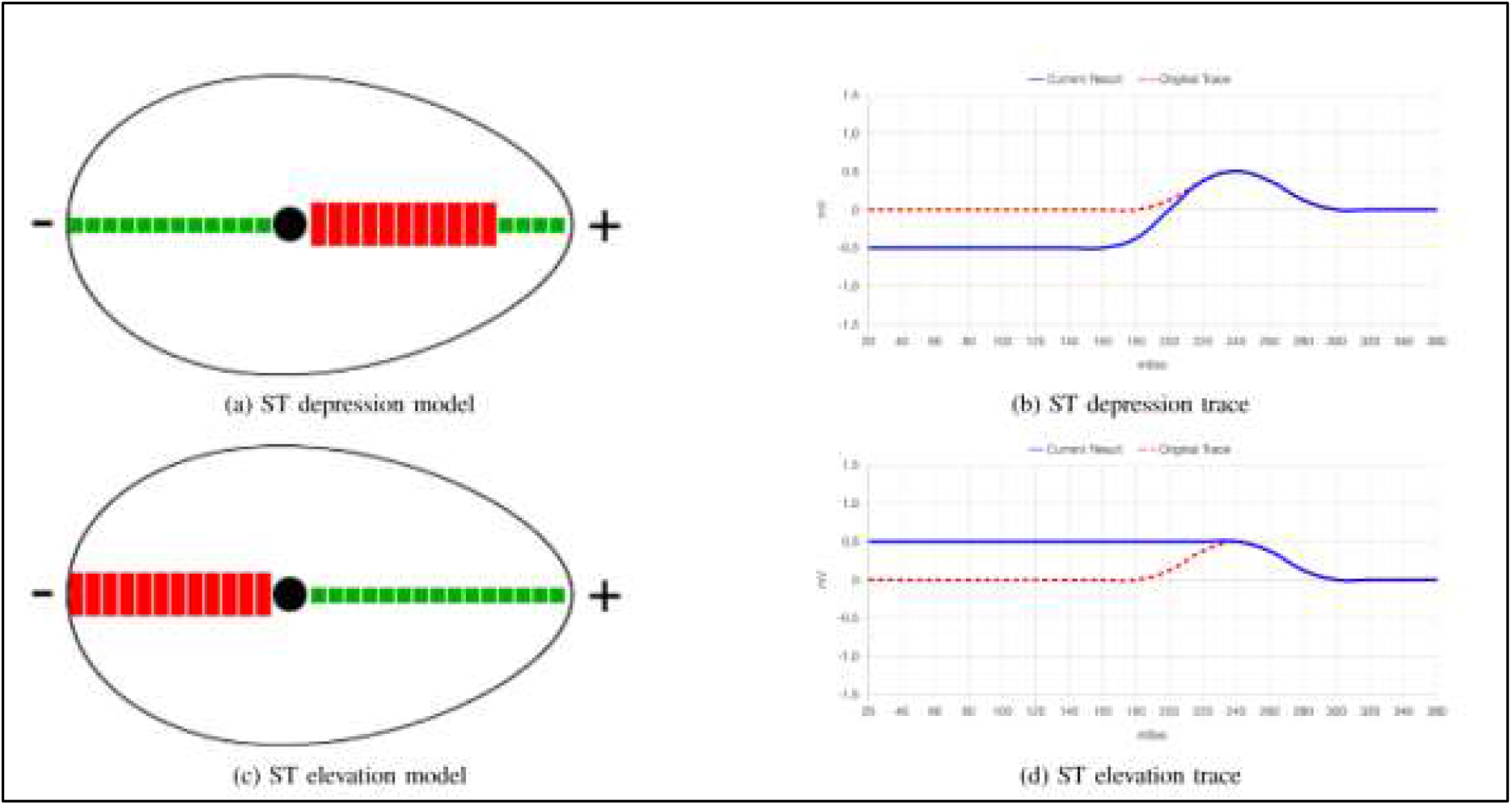
ST depression and ST elevation models and traces. The diagrams illustrate how ST depression and ST elevation traces on an ECG can result from the Simplified Egg and Changing Yolk Model.

In Figure 5a, as the repolarisation signal spreads, there is a deficit of positive deflection and this results in ST depression because the signal seems to be moving away from the lead overall. Since the damage is close to the WCT, the ST depression occurs right from the start of repolarisation. If the damaged region of the heart is situated at a greater distance from the WCT, the resulting ST depression will manifest later. Consequently, the onset of ST depression signifies the location of damage relative to the WCT and the direction of the lead. In Figure 5c, the damage is on the opposite side of the WCT, which results in ST elevation in this case because of the lack of transmission in negative deflection.

The proposed model can also be used to investigate other abnormal T-wave morphologies. The sequence in Figure 6 represents the regions on the lead which register the damaged areas and their resulting traces. They depict the commonly seen abnormal T-wave morphologies of higher T-waves, flattened T-waves, inverted T-waves, and biphasic T-waves along with drawings of their registered damaged areas within the egg shape.

**Figure 6:**
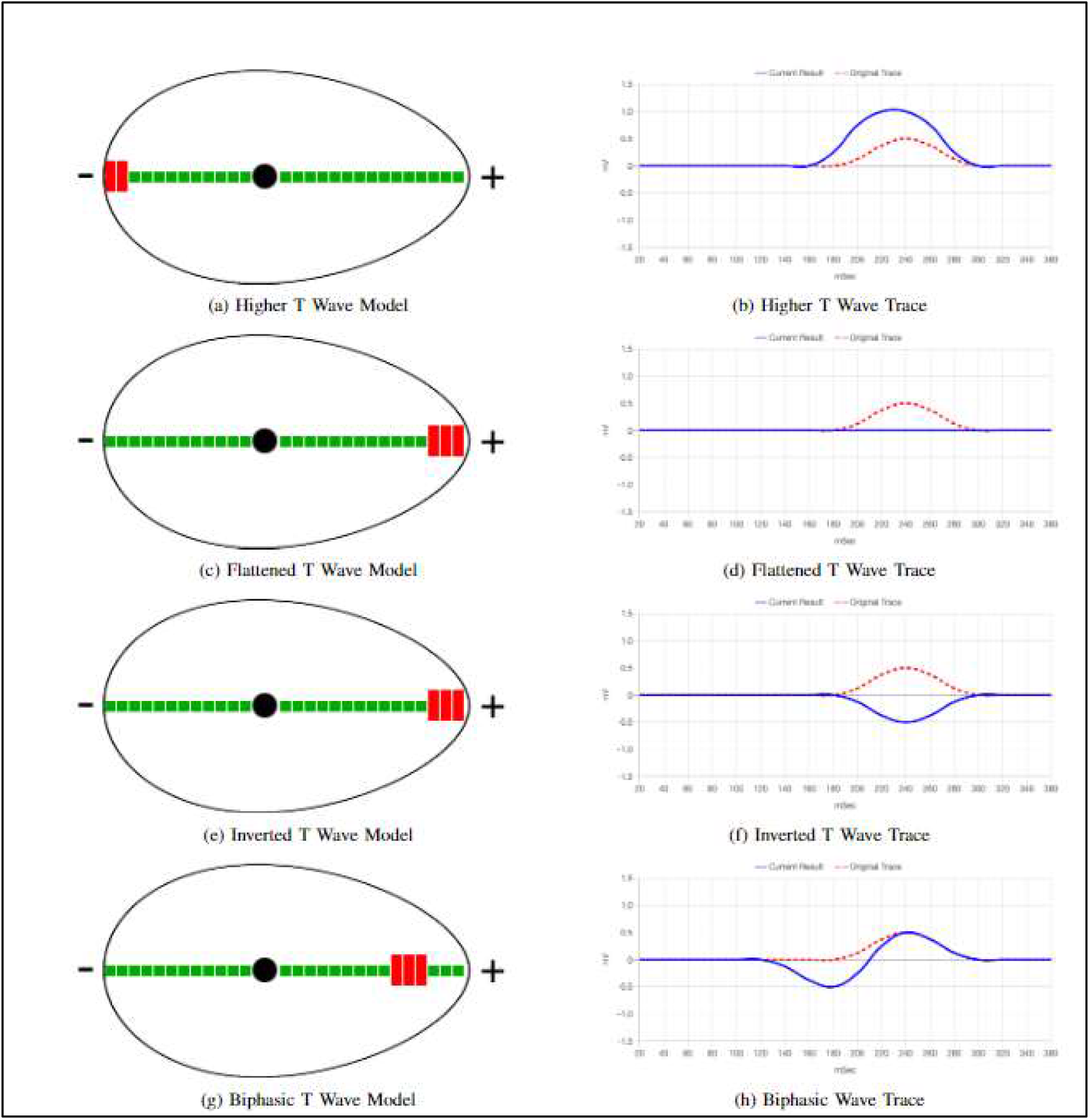
Representation of Abnormal T-wave Morphologies on ECG Traces. These diagrams demonstrate how the Simplified Egg and Changing Yolk Model can reproduce four common abnormal T-wave morphologies on an ECG by placing damaged registered areas (red blocks) in specific parts of the egg shape.

In Figure 6a, the higher T-wave is depicted as resulting from muscle damage on the negative side of the WCT for the lead. However, other factors such as hypertrophy and ion imbalance, can also produce higher T-waves. Note that while the settings for the flattened and inverted T-waves (Figures 6c and 6e) seem similar, they differ in hypertrophy and damage severity to reproduce the T-wave morphologies. Further details on these examples can be explored using the webtool available at: https://t-wave.aber.ac.uk/

A comprehensive range of T-wave shapes, along with corresponding real-world ECG examples can illustrate how this simple, yet versatile model can emulate various real-life T-wave abnormalities and explain the presence of three distinct T-wave shapes (positive, biphasic, and inverted) following ST depression, compared to the single T-wave shape (positive) observed after ST elevation in lead V5. The proposed model also demonstrates how the model can account for variations in T-wave shapes in ‘normal ECGs’, depending on the orientation and shape of the heart relative to lead direction (see Supplementary Material section 2).

## Discussion

We have demonstrated that the novel Simplified Egg and Changing Yolk Model is a multifaceted model for illustrating the conditions that give rise to various T-wave morphologies in ECG traces. The proposed model specifically applies to the repolarisation process, with the T-wave representing ventricular repolarisation. Since ventricular depolarisation travels quickly through cardiac nerve fibres, like the bundle of His, bundle branches, and Purkinje fibres, necessitating a different model. In contrast, the ventricular repolarisation process, which resets the depolarised ventricular muscles, is not facilitated by these nerve fibres, making the Simplified Egg and Changing Yolk Model suitable for replicating this process. While the examples used lead V5, these principles apply to other ECG leads as well. However, the orientation of each lead within the ‘egg shape’ must be considered. Lead V5 was chosen for demonstration due to its alignment with the heart apex, matching the most asymmetric part of the ‘egg shape’.

The implications of this model of ventricular repolarisation challenges the conventional view that the repolarisation process commences only at the conclusion of the S wave. Instead, it suggests repolarisation starts with the Q-wave as depolarisation begins and continues as a single process through to the end of the T-wave. Consequently, the ST interval and T-wave interval should be considered as a unified process, rather than being divided into separate components as is currently believed^16^. Secondly, given that ST depression in one lead manifests as ST elevation in its reciprocal lead, it becomes difficult to reliably interpret ST depression as indicative of myocardial ischaemia or ST elevation as representative of myocardial infarction^17^. Using this model, it is more appropriate to consider the entire repolarisation process as a single unit and attempt to determine ‘where’ the myocardial issue is occurring (rather than focusing on ‘what’ is happening). Other investigative methods, such as cardiac enzyme analysis, should be used as markers for potential myocardial damage. Thirdly, there are implications for use of the QTc interval. Currently the QTc interval ends when repolarisation exits the ‘T-zone’ of the Simplified Egg and Changing Yolk Model. This approach uses the ‘front’ of the wave of the repolarisation signal (Q-wave start) and the ‘tail end’ of the wave of the repolarisation signal when it exits the ‘T-zone’. In practice, challenges arise in calculating the QTc interval when a U-wave appears before the T-wave ends or when a P-wave begins before the T-wave concludes. For examples of these challenges in QTc calculation, please refer to Supplementary Material section 1.

There is no universally accepted formula for calculating the QTc interval. Multiple recognised formulae exist, each with advantages and limitations, leading to potential discrepancies in clinical practice^18^. The most commonly used are Bazett’s, Fridericia’s, Hodges’ and Framingham’s formulae. The choice of the most appropriate formula should be carefully considered based on the clinical context and population studied^19^. The proposed model suggests measuring the interval from the onset of the Q-wave up to the T-wave peak, termed the QTp interval. This interval represents the duration from the start of repolarisation at the Q-wave to the outer boundary of the ‘T-zone’ at the T-wave peak. Measuring the QTp interval reduces ambiguity in the QT interval measurement and simplifies the automated detection of the T-wave peak. This is contrary to calculating the T-wave end based on the different formulae used by the ECG machine and this approach is supported by multiple studies^20–22^. For biphasic T-waves, the highest T-wave peak should be used, and for inverted T-waves the lowest trough position is used. However, in cases of flat T-waves, neither the QTp nor the QTc interval can be determined. The proposed model lends stronger support to the case for using the QTp interval as a more reliable and accurate parameter, especially in assessing the risk of arrhythmia caused by potential ‘R on T phenomenon’, and in measuring the effects of medication on the QT interval^23–25^.

Finally, the Simplified Egg and Changing Yolk Model can be used to reproduce various ECG abnormalities, including ST elevation, ST depression and T-wave flattening or inversion. By selecting multiple blocks to simulate “damage” in the model, one can reflect the distribution of the affected artery. Future investigations could modify the model to incorporate coronary artery distribution, aiming to replicate the effects of occlusion in specific arteries.

## Conclusion

In conclusion, this study introduces the Simplified Egg and Changing Yolk Model, a comprehensive framework that enhances our understanding of T-wave morphologies on ECGs during ventricular repolarisation. By offering a unified model that explains various T-wave patterns, this work could improve ECG interpretation and lead to more accurate diagnoses of cardiac abnormalities.

Furthermore, the proposed model underscores the limitations of the widely used QTc interval in assessing arrhythmia risk and supports the QTp interval as a more reliable alternative. By demonstrating the QTp interval’s effectiveness in evaluating arrhythmia risk and the impact of QT-prolonging medications, this model offers a valuable tool for improving risk stratification and patient management in cardiac diseases. We recommend adopting the more reliable QTp (Q-wave start to T-wave peak) interval over the QTc interval, especially for assessing medication effects on the QT interval.

Our work has important implications for clinical practice and research, offering a better understanding of ventricular repolarisation on ECGs. This can improve decision-making in patient management and aid in developing new therapeutic strategies. Our recommendation for the QTp interval paves the way for future research to validate its utility across different populations and settings and to refine the Simplified Egg and Changing Yolk Model. This study represents a significant step toward improving the assessment and management of cardiac conditions, with the potential to enhance patient outcomes and overall cardiovascular health.

Building on this work, we plan to explore the ventricular repolarisation signal’s behaviour as it propagates beyond the T-zone within the egg shape, potentially uncovering the origin of the U-wave. We also aim to apply this model to the atrial repolarisation process and explore its broader implications. By expanding our insights, we can advance understanding in cardiac electrophysiology, leading to improved patient care and outcomes.

## Supporting information

Supplementary Materials for Deciphering T wave Morphologies on ECGs

## Data Availability

The study used ECG traces from PhysioNet.
Reference:
Perez Alday EA, Gu A, Shah A, et al.: Classification of 12-lead ECGs: The PhysioNet/Computing in Cardiology Challenge 2020 [Internet]. PhysioNet, [cited 2024 Dec 11],. Available from: https://physionet.org/content/challenge-2020/1.0.2/

## References

1. Kaplan Berkaya S, Uysal AK, Sora Gunal E, Ergin S, Gunal S, Gulmezoglu MB: A survey on ECG analysis. Biomedical Signal Processing and Control 2018; 43:216– 235.

2. Kligfield P, Gettes LS, Bailey JJ, et al.: Recommendations for the Standardization and Interpretation of the Electrocardiogram: Part I: The Electrocardiogram and Its Technology: A Scientific Statement From the American Heart Association Electrocardiography and Arrhythmias Committee, Council on Clinical Cardiology; the American College of Cardiology Foundation; and the Heart Rhythm Society Endorsed by the International Society for Computerized Electrocardiology. Circulation 2007; 115:1306–1324.

3. Nielsen JC, Lin Y-J, De Oliveira Figueiredo MJ, et al.: European Heart Rhythm Association (EHRA)/Heart Rhythm Society (HRS)/Asia Pacific Heart Rhythm Society (APHRS)/Latin American Heart Rhythm Society (LAHRS) expert consensus on risk assessment in cardiac arrhythmias: use the right tool for the right outcome, in the right population. EP Europace 2020; 22:1147–1148.

4. Hammad M, Maher A, Wang K, Jiang F, Amrani M: Detection of abnormal heart conditions based on characteristics of ECG signals. Measurement 2018; 125:634– 644.

5. Savonitto S: Prognostic Value of the Admission Electrocardiogram in Acute Coronary Syndromes. JAMA 1999; 281:707.

6. Katz AM: T Wave “Memory”: Possible Causal Relationship to Stress-Induced Changes in Cardiac Ion Channels? Cardiovasc electrophysiol 1992; 3:150–159.

7. Cardona A, Zareba KM, Nagaraja HN, et al.: T-Wave Abnormality as Electrocardiographic Signature of Myocardial Edema in Non-ST-Elevation Acute Coronary Syndromes. JAHA 2018; 7:e007118.

8. Klabunde RE: Cardiovascular physiology concepts. 2. ed. Philadelphia: Wolters Kluwer [u.a.], 2012,.

9. Sagie A, Larson MG, Goldberg RJ, Bengtson JR, Levy D: An improved method for adjusting the QT interval for heart rate (the Framingham Heart Study). The American Journal of Cardiology 1992; 70:797–801.

10. Goldenberg I, Moss AJ, Zareba W: QT Interval: How to Measure It and What Is “Normal.” Cardiovasc electrophysiol 2006; 17:333–336.

11. Panikkath R, Reinier K, Uy-Evanado A, et al.: Prolonged Tpeak-to-Tend Interval on the Resting ECG Is Associated With Increased Risk of Sudden Cardiac Death. Circ: Arrhythmia and Electrophysiology 2011; 4:441–447.

12. Al-Khatib SM, LaPointe NMA, Kramer JM, Califf RM: What Clinicians Should Know About the QT Interval. JAMA [Internet] 2003 [cited 2024 Dec 10]; 289. Available from: http://jama.jamanetwork.com/article.aspx?doi=10.1001/jama.289.16.2120

13. Giudicessi JR, Noseworthy PA, Ackerman MJ: The QT Interval: An Emerging Vital Sign for the Precision Medicine Era? Circulation 2019; 139:2711–2713.

14. Wilson FN, MacLeod AG, Barker PS: The T deflection of the electrocardiogram. Trans Assoc Am Physicians 46:29–38.

15. Meijborg VMF, Conrath CE, Opthof T, Belterman CNW, De Bakker JMT, Coronel R: Electrocardiographic T Wave and its Relation With Ventricular Repolarization Along Major Anatomical Axes. Circ: Arrhythmia and Electrophysiology 2014; 7:524–531.

16. Goldberger AL, Shvilkin A, Goldberger ZD, Goldberger AL: Goldberger’s clinical electrocardiography: a simplified approach. Ninth Edition. Philadelphia, PA: Elsevier, 2017,.

17. Pollehn T, Brady WJ, Perron AD, Morris F: The electrocardiographic differential diagnosis of ST segment depression. Emergency Medicine Journal 2002; 19:129– 135.

18. Vandenberk B, Vandael E, Robyns T, et al.: Which QT Correction Formulae to Use for QT Monitoring? JAHA 2016; 5:e003264.

19. Suárez-León AA, Varon C, Willems R, Van Huffel S, Vázquez-Seisdedos CR: T-wave end detection using neural networks and Support Vector Machines. Computers in Biology and Medicine 2018; 96:116–127.

20. Madeiro JPV, Nicolson WB, Cortez PC, et al.: New approach for T-wave peak detection and T-wave end location in 12-lead paced ECG signals based on a mathematical model. Medical Engineering & Physics 2013; 35:1105–1115.

21. Schläpfer J, Wellens HJ: Computer-Interpreted Electrocardiograms. Journal of the American College of Cardiology 2017; 70:1183–1192.

22. Postema P, Wilde A: The Measurement of the QT Interval. CCR 2014; 10:287–294.

23. Liu MB, Vandersickel N, Panfilov AV, Qu Z: R-From-T as a Common Mechanism of Arrhythmia Initiation in Long QT Syndromes. Circ: Arrhythmia and Electrophysiology 2019; 12:e007571.

24. Sundqvist K, Sylvén C: Cardiac repolarization properties during standardized exercise test as studied by QT, QT peak and terminated T-wave intervals. Clinical Physiology 1989; 9:419–425.

25. Haapalahti P, Viitasalo M, Perhonen M, et al.: Comparison of QT peak and QT end interval responses to autonomic adaptation in asymptomatic LQT1 mutation carriers: QT peak versus QT end responses in LQT1. Clinical Physiology and Functional Imaging 2011; 31:209–214.

