## Supplementary Materials for Deciphering T wave Morphologies on ECGs for "Deciphering T-wave Morphologies on ECGs: The Simplified Egg and Changing Yolk Model and the Importance of the QTp Interval"

#### Section 1: QTc Interval Analysis

##### 1.1 Introduction

The following supplementary material delves into key concepts and challenges of the QTc interval, which adjusts the time between the start of the Q wave and the end of the T wave for heart rate, serving as a crucial biomarker for cardiovascular health. Furthermore, it delves into the nuances involved in QTc calculation, which often pose difficulties in both clinical and research settings. This supplementary information is structured into two main parts. The first part, "Complexities and Challenges in QTc Interval Analysis: Enhancing Heart Rate Variability Assessments," provides a deeper understanding of physiological variations and their impact on QTc intervals. Here, we address potential measurement errors due to U waves, disparities caused by regional repolarisation differences, and the critical aspects of inter-lead variability. Additionally, we elaborate on the pressing need for a more uniform and robust method of standardisation within this analysis. The aim of this part is to emphasise the importance of comprehensive QTc analysis, particularly considering its relevance to heart rate variability assessments.

The second part, "Key Challenges Associated with QTc Calculation," focuses on the intricate task of calculating the QTc interval accurately. The sections are devoted to explicating the complex relationship between heart rate and QTc calculation and highlighting the complexities of QT interval identification. An overview is provided on the implications of drug-induced QTc prolongation, the influence of electrolyte imbalances, and the impact of specific disease states on the QTc interval. This part intends to present a complete picture of the challenges associated with the calculation of QTc and the influencing factors.

Collectively, this supplementary material aims to provide an in-depth view of the complexities and challenges associated with QTc interval analysis and calculation. The objective is to stimulate further research, encourage dialogue on standardisation, and improve methodologies for a more accurate understanding of heart rate variability, which plays a pivotal role in cardiovascular health. We hope that this additional information assists readers in understanding the nuances of QTc interval analysis and its calculation, thereby enhancing the main findings and implications of our research paper.

#### 1.2 Part 1. Complexities and Challenges in QTc Interval Analysis: Enhancing Heart Rate Variability Assessments

The corrected QT interval (QTc) represents the duration from the start of the Q wave to the end of the T wave in an electrocardiogram (ECG), a time span that signifies the total time taken for ventricular depolarisation and repolarisation. Clinically, it is a vital parameter to measure as it can reveal potential cardiac arrhythmias. However, using QTc interval for comparing heart rate variability presents several challenges. These include representing physiological changes within patients, addressing measurement errors caused by U waves superimposing on the T wave, accounting for regional repolarisation differences within an individual's heart, and managing inter-lead variability along with a consensus on a standard lead to use.

##### 1.2.1 Physiological Variations and QTc interval

The QTc interval is sensitive to multiple physiological factors. These include autonomic nervous system activity[1], metabolic changes[2], circadian rhythms[3], and physical activity[4] among others. Each of these factors can significantly impact the heart rate, and consequently the QTc interval. For instance, during periods of physical exercise or stress, the activity of the sympathetic nervous system increases, leading to a higher heart rate. This increase in heart rate usually shortens the QTc interval. Conversely, during periods of rest or sleep, when the parasympathetic nervous system is more active, the heart rate decreases, typically resulting in a lengthened QTc interval. Metabolic changes, such as hypoglycemia or hyperglycemia, can also influence the QTc interval. Both these states can lead to changes in the electrophysiological activity of the heart, potentially altering the QTc duration.

Similarly, the body's internal biological clock, or circadian rhythm, also has a profound impact on heart rate variability. The heart rate and QTc interval have been found to exhibit diurnal variations, with changes occurring across the day and night.

Given this sensitivity to physiological changes, it's crucial to normalise these variables when comparing QTc intervals among different cases. For example, comparing the QTc interval of a person at rest with someone who has just finished exercising could lead to incorrect conclusions due to the significant physiological differences between the two states. Therefore, accurate comparisons necessitate that the physiological state at the time of QTc measurement be similar or taken into account during interpretation[5].

##### 1.2.2 Measurement Errors Due to U waves

U waves are a part of the ECG that follow the T wave and precede the next P wave. They are generally small, often not exceeding 1-2 mm in height on the ECG, and their origin is still a matter of ongoing research. One of the common challenges in the measurement of the QTc interval is the potential overlap of the U wave with the T wave, which may cause an overestimation of the QTc interval duration[6]. This superimposition can occur due to several reasons, such as slower heart rates, electrolyte imbalances like hypokalemia, or the use of certain medications. When the U wave overlaps with the T wave, it extends the apparent repolarisation time leading to an overestimation of the QTc interval. This can be particularly problematic in situations where a precise measurement of the QT interval is critical, such as in the diagnosis of Long QT Syndrome.

To overcome this challenge, it is important to enhance the ECG analysis algorithms to detect and account for U waves. Advanced signal processing techniques, including wavelet transformation and machine learning algorithms, could be employed to more accurately differentiate T waves from superimposed U waves. This would allow for a more precise measurement of the QTc interval and a better comparison of heart rate variability between different cases.

##### 1.2.3 Regional Repolarisation Differences

The human heart, despite being a single organ, possesses intricate regional differences in its electrophysiological activity. These differences are more prominent during the repolarisation phase, represented by the T wave on the ECG. The duration and shape of the T wave, and hence the QTc interval, may vary across different regions of the heart, based on factors like myocardial thickness, cell type, and local autonomic innervation. Regional differences in repolarisation times can result from several physiological and pathological conditions[7,8]. For example, conditions like myocardial ischemia or infarction, electrolyte imbalances, or genetic disorders can create regional heterogeneity in the heart's electrical activity. This variability is usually reflected in the ECG as T wave alterations. However, when using the QTc interval to compare heart rate variability, these regional differences can pose a challenge. Since the ECG leads capture the electrical activity from different angles, this can result in varying QTc intervals, depending on the repolarisation time of the specific region viewed by the lead[9].

To address this, innovative techniques are required that can account for the regional differences in repolarisation times. This might involve using multiple leads to capture a more global measure of repolarisation or incorporating advanced computational models that can predict the regional variations in QTc intervals based on individual characteristics.

##### 1.2.4 Inter-Lead Variability and Standardisation

In an ECG, each lead offers a unique 'view' of the heart's electrical activity, owing to its specific orientation with respect to the heart. Therefore, the QT interval recorded can vary significantly from one lead to another. This variation, termed as inter-lead variability, can introduce potential inaccuracies when comparing QTc intervals among different cases or even within the same individual over time if different leads are used.

Certain leads, such as the precordial leads V2-V3, are known to display longer QT intervals, whereas others like lead I and aVL often show shorter intervals[10]. Furthermore, the presence of cardiac pathologies or even normal anatomical variations can amplify these differences. Due to this inherent variability, the choice of lead used for measuring the QTc interval becomes critically important. However, there is a lack of consensus on a standard lead to use for QTc measurement, which can confound comparisons of heart rate variability[11].

To address this, it is essential to establish a standardised protocol for the selection of the lead for QTc measurement. This might involve selecting the lead that best represents the global QTc interval or consistently using the same lead for all measurements. Furthermore, the development of multilead or vectorcardiographic approaches, which consider the information from multiple leads simultaneously, could offer a more comprehensive view of the QTc interval and help overcome the limitations posed by inter-lead variability.

#### 1.3 Part 2. Key challenges associated with QTc calculation.

##### 1.3.1 Heart Rate and QTc Calculation

The QT interval represents the time taken for both ventricular depolarisation and repolarisation. It is measured from the beginning of the QRS complex to the end of the T wave on the ECG. However, the raw QT interval is affected by heart rate - it shortens as the heart rate increases and lengthens as the heart rate decreases. Thus, to allow comparisons between different heart rates, a corrected QT interval (QTc) is used.

Various correction formulas have been proposed over the years to adjust the QT interval for heart rate, with the most commonly used being Bazett's, Fridericia's, Framingham's, and Hodges' formulas. Bazett's formula, the most commonly used, is defined as $QTc = QT/\sqrt{RR}$, where RR is the interval from the onset of one QRS complex to the onset of the next, measured in seconds. Despite its widespread use, Bazett's formula tends to overcorrect at high heart rates and undercorrect at low heart rates, leading to potential inaccuracies[4]. Fridericia's formula, defined as $QTc = QT/RR^{1/3}$, and Framingham's formula, defined as $QTc = QT + 0.154*(1-RR)$, have been proposed as alternatives to address the shortcomings of Bazett's formula. Hodges' formula, defined as $QTc = QT + 1.75*(\text{heart rate} - 60)$, is another alternative that is considered to be more accurate across different heart rates[12].

Despite these various correction methods, none are perfect, and each has its limitations. For example, all these formulas are based on population averages and may not accurately correct the QT interval in individual cases. Moreover, these corrections are derived from resting ECGs and may not apply during exercise or other states of increased heart rate. Thus, QTc calculation remains a complex task, especially in situations with significant heart rate variability[13].

##### 1.3.2 QT Interval Identification

QT interval measurement is the basis for calculating the QTc. It begins with the start of the Q wave and ends at the end of the T wave on the ECG. However, accurate identification of these points can sometimes be challenging due to a number of factors.

Firstly, the Q wave can sometimes be small and difficult to distinguish, particularly in certain ECG leads. This can lead to variability in identifying the start of the QT interval.

Secondly, defining the end of the T wave, and hence the QT interval, can often be tricky. The T wave end is generally defined as the intersection of the tangent to the steepest downslope of the T wave and the baseline. However, in certain cases, the T wave can be low in amplitude, biphasic, or can merge with a U wave, making it challenging to accurately identify its end. Improper identification of the Q wave start or T wave end can lead to significant errors in the calculated QTc interval. These errors can potentially result in misdiagnosis or inaccurate risk stratification. To minimise this, automated computer algorithms are often used. However, these too can make errors, and manual checking by a trained individual remains the gold standard.

##### 1.3.3 Drug-induced QTc Prolongation

Certain medications can interfere with the cardiac electrical activity by inhibiting specific ion channels involved in ventricular repolarisation, notably the hERG (human Ether-à-go-go-Related Gene) potassium channel. This channel blockade leads to a delay in the repolarization process, extending the duration of the QT interval, seen as QTc prolongation on an ECG. A prolonged QTc due to medications is of significant clinical concern, as it raises the risk of a severe form of ventricular tachycardia called Torsades de Pointes, which can be potentially life-threatening. This necessitates careful QTc monitoring in patients on such medications[14].

However, the pharmacological impact of these drugs adds a layer of complexity when calculating the QTc interval. When a person is on QT-prolonging drugs, the measured QTc interval is not just a reflection of their inherent QTc but also encapsulates the drug's effect.

Interpretation in such instances requires consideration of the drug's influence and timing. Understanding the pharmacokinetics of the drug, for instance, can assist in identifying when its effect on QTc is at its peak. In certain cases, under the supervision of a healthcare provider, it might be required to adjust or even momentarily stop the medication to ascertain an accurate measure of the individual's intrinsic QTc interval.

##### 1.3.4 Electrolyte Imbalances

Electrolytes, primarily potassium, calcium, and magnesium, play critical roles in the electrical activities of the heart, particularly in the processes of depolarisation and repolarisation. These ions move in and out of cardiac cells through specific channels, generating electrical signals that regulate the heartbeat. When there's an imbalance in the concentration of these electrolytes, the heart's normal electrical activity can be disrupted, leading to changes in the QT interval on the ECG[15].

*Potassium:* This electrolyte plays a pivotal role in repolarisation, the phase represented by the QT interval. Low levels of potassium in the blood, or hypokalemia, can delay repolarisation, leading to prolongation of the QT interval. On the other hand, high potassium levels, or hyperkalemia, can shorten the QT interval.

*Calcium:* Calcium is involved in the depolarisation phase of the heart's electrical activity. High levels of calcium, or hypercalcemia, can shorten the QT interval, while low calcium levels, or hypocalcemia, are usually associated with a prolonged QT interval.

*Magnesium:* Magnesium acts as a natural calcium blocker and plays a critical role in maintaining potassium balance. Low levels of magnesium, or hypomagnesemia, can lead to both potassium loss and prolongation of the QT interval[16].

In patients with electrolyte imbalances, measuring and interpreting the QTc interval can be challenging due to these effects. It is crucial to consider the patient's electrolyte status, and in some cases, it may be necessary to correct the electrolyte imbalances before an accurate QTc measurement can be obtained. Additionally, long-term management should involve treating the underlying cause of the imbalance to prevent further cardiac complications.

##### 1.3.5 Disease States

Underlying disease states can significantly impact the heart's electrical activity, including the QTc interval. These effects can stem from direct influences on the heart or indirect effects on the systems that regulate heart function.

*heart diseases:* Conditions such as coronary artery disease, congestive heart failure, and cardiomyopathies can affect the QTc interval. These diseases lead to structural and functional changes in the heart muscle that can alter the timing of ventricular repolarization, thereby affecting the QTc interval[17]. For instance, ischemic heart disease, resulting from reduced blood flow to the heart muscle, can prolong the QT interval.

*Liver diseases:* Advanced liver diseases, such as cirrhosis, can lead to QTc prolongation. The link between liver disease and QTc prolongation isn't fully understood, but it is thought to involve several factors, including autonomic dysfunction and alterations in cardiac ion channels due to cirrhosis. Liver disease can also affect electrolyte balance, further influencing the QTc interval[18].

*Endocrine disorders*: Disorders of the endocrine system, particularly thyroid disorders, can affect the QTc interval. Hypothyroidism can cause a lengthening of the QTc interval due to decreased metabolic rate and associated changes in heart function. Conversely, hyperthyroidism can result in QTc shortening due to increased metabolism and heart rate, although QTc prolongation may also occur.

*Neurological disorders:* Certain neurological conditions, such as autonomic neuropathy, can also affect the QTc interval by altering the balance of the autonomic nervous system, which regulates heart rate and rhythm[19].

##### 1.4 Conclusion

In conclusion, this supplementary material has provided an extensive discussion on the plethora of challenges and complexities associated with defining the QTc interval in an ECG. The QTc interval's paramount role in risk stratification for developing arrhythmias underscores the urgency to address these problems. The quest for alternatives must prioritise not just their practicality and reliability but also their robust grounding in the underlying physiology.

The main paper ventures a proposition that QTp could serve as a more efficient and precise measure compared to the traditional QTc interval. As we continue to grapple with the challenges of QTc interval analysis and calculation, it is the diligent examination of such alternatives that will guide the path forward in our endeavour to enhance cardiovascular health assessments.

We believe this supplementary material serves as a detailed backdrop to the main paper, underlining the significance of the challenges in QTc analysis and calculation, and thus the potential impact of shifting to QTp as a measurement tool. We hope that the insights provided in this document offer a holistic understanding of the complexities involved and thus underscore the need for rigorous and continuous research in this area of cardiovascular health.

#### Section 2: Evidence for the Simplified Egg and Changing Yolk Model Usefulness in the Real-World Context

##### 2.1 Introduction

This supplementary section uses ECG examples from the publicly available PTB-XL dataset[20] to provide proof as to the usefulness of the Simplified Egg and Changing Yolk Model in explaining the mechanism behind the various T wave (ventricular) repolarisation trace shapes.

Although the PTB-XL dataset itself contains 21,837 ECGs, this supplementary paper gives ECG examples demonstrating the various T wave configuration in lead V5 using the first 1000 ECGs (training group g1) of the dataset.

This section has been presented in three parts.

The **first part** demonstrates how the Simplified Egg and Changing Yolk model can be used to emulate the following T wave shapes:

- Example 1. Normal T wave
- Example 2. Higher (tented) T wave
- Example 3. Flat (pathology related) T wave
- Example 4. Inverted T waves
- Example 5. Biphasic T wave with initial positive deflection
- Example 6. Biphasic T wave with initial negative deflection
- Example 7. Bifid (camel hump shaped) T wave

The Simplified Egg and Changing Yolk model proposes that the ventricular repolarisation trace can be thought of as a signal propagating from the WCT, and the models explain how the ST segment is formed by symmetrical part of the egg shape, whilst the T wave if formed by the asymmetrical part.

Inevitably with a simplified model there are trade-offs to be made in terms of how closely the simulated T waves match the ECG ventricular repolarisation patterns. This we see especially in matching the Bifid T wave (and the ST depression followed by a negative or inverted T wave in the next part). The model can be improved in several ways however for clarity in explaining the concept of how T wave repolarisation shapes occur we have left the model in its simplified form.

The **second part** demonstrates how the Simplified Egg and Changing Yolk Model can be used to emulate:

Example 8. ST depression followed by positive T wave

Example 9. ST depression followed by biphasic T wave

Example 10. ST depression followed by inverted T wave

Example 11. ST elevation followed by positive T wave

Because of the asymmetrical nature of the ‘egg’ shape in the model, the Simplified Egg and Changing Yolk model predicts that it would be difficult to see ST elevation followed by specific configurations of T wave: biphasic and inverted. Thus, although we see ‘ST depression’ followed by T waves that are positive, biphasic or inverted, in ‘ST elevation’ we would only see ST elevation followed by a positive T wave. This is in keeping with what is seen in real world ECG examples providing further evidence of the usefulness of this model.

The **third part** demonstrates how the Simplified Egg and Changing Yolk model provides an explanation as to why ‘normal’ T waves on an ECG have a different appearance depending on the heart rotation and orientation. In particular when the ‘egg shape’ from the lead V5 point of view appears to be:

Example 12. Asymmetrical – Normal T wave following a baseline ST segment

Example 13. Slightly asymmetrical – Normal T wave that start soon after QRS wave

Example 14. Symmetrical – Flat non-pathological T wave

We conclude that the simple but versatile Simplified Egg and Changing Yolk model is useful in understanding how the ventricular repolarisation pattern can lead to the various T wave traces on an ECG. This is done here by looking specifically at the direction of lead V5 but is applicable to the other ECG leads depending on the ‘shape’ of the heart they are seeing.

In the future we plan to further refine and develop this model to investigate the explanation for the U wave appearance, and also how different types of hypertrophy and coronary artery territory damage affects the shape of the subsequent repolarising T wave.

#### 2.2 Part 1: Variations of the T Wave Shapes

In Examples 1 to 7 below we describe how the Simplified Egg and Changing Yolk model can be used to describe the ‘normal’ T wave shape, and many of the common variations of T wave shapes due to pathological causes.

Real world ECG examples from the PTB-XL dataset are given alongside each T wave morphology example.

These examples can be explored in more detail using our software simulation tool available at: https://t-wave.aber.ac.uk/

#### Example 1. Normal T Wave

**ECG ID: training/ptb-xl/g1/HR00029**

**ECG Diagnosis Given:** sinus rhythm normal ecg.

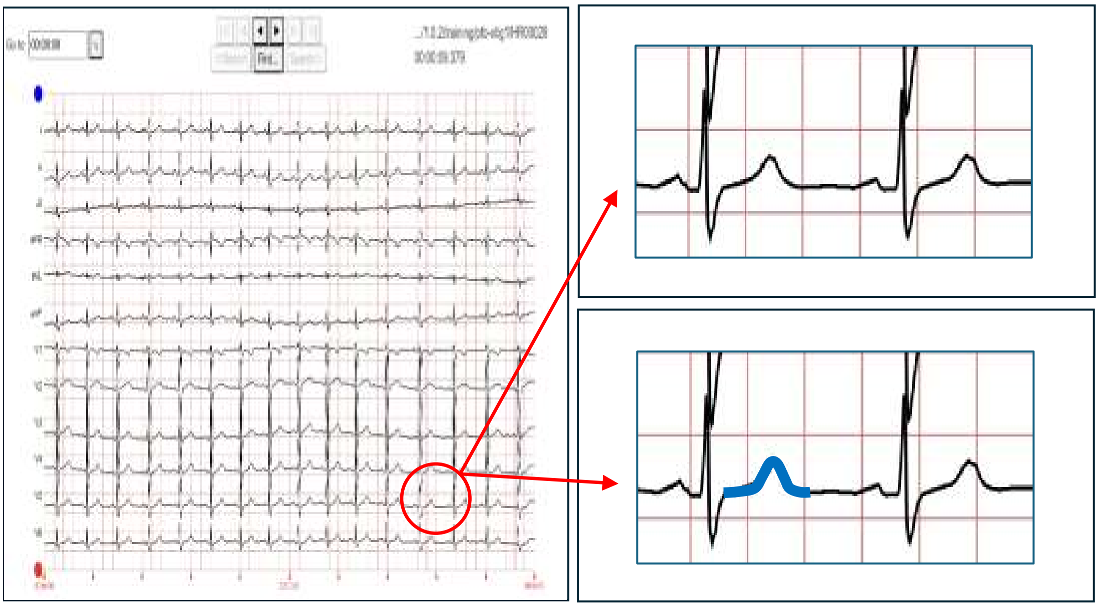

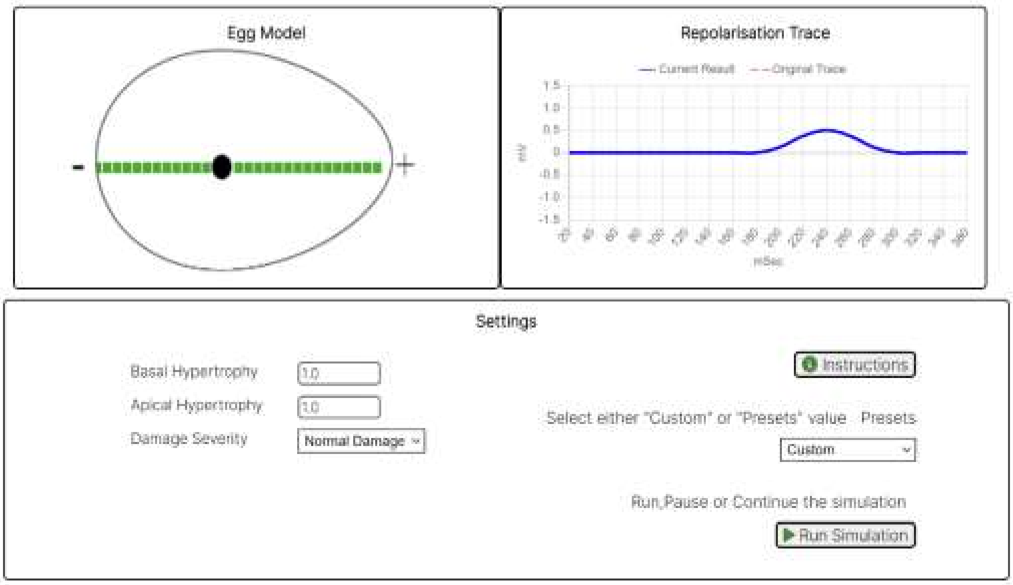

**Commentary**: This is typical example of a normal repolarisation trace in lead V5. The ST segment of the simulated trace is generated by the symmetrical part of the egg shape, and the T wave is generated when the signal propagates into the asymmetrical part.

**Other Similar Example Cases:** training/ptb-xl/g1/HR00054, HR00120, HR00198, HR00322, HR00488, HR00507, HR00644, HR00726, HR00824, HR00949

291

#### Example 2. Higher T Wave (Tented Shape)

**ECG ID: training/ptb-xl/g1/HR00521**

**ECG Diagnosis Given:** sinus rhythm. non-specific increased st-t changes otherwise normal ecg.

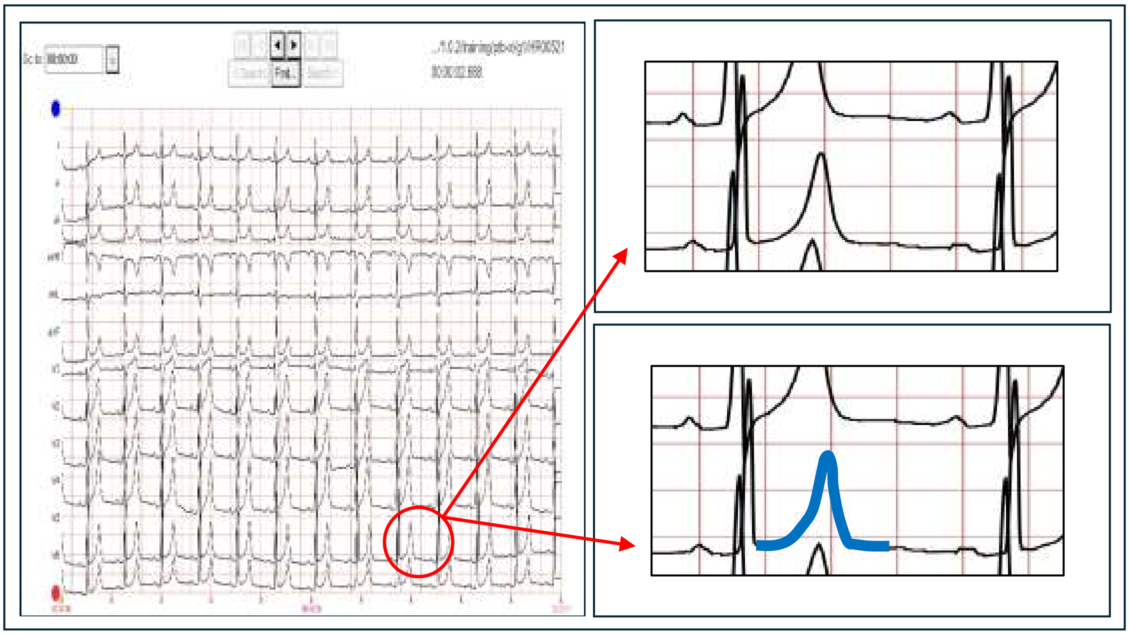

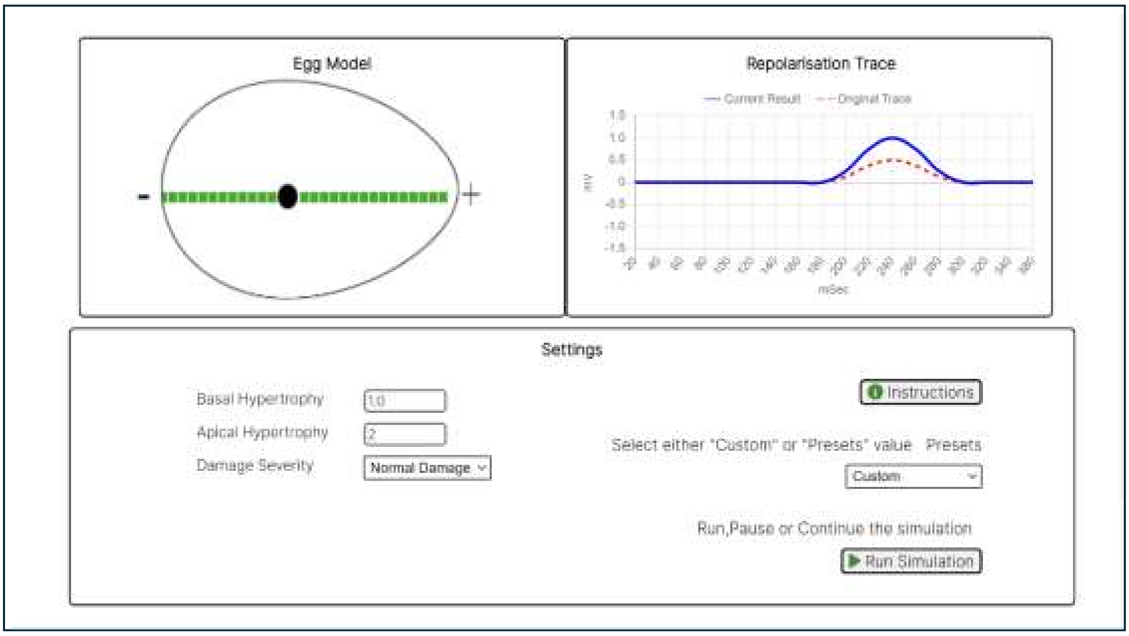

**Commentary**: This is an example of a prominent T wave repolarisation trace in lead V5. Tall T waves can appear in acute coronary ischaemia / infarction, pericarditis, or hyperkalaemia. They can also be present in some adults and athletes.

**Other Similar Example Cases:** training/ptb-xl/g1/HR00306, HR00682, HR00718, HR00821, HR00974

292

#### Example 3. Flat T Wave (Pathological)

**ECG ID: training/ptb-xl/g1/HR00476**

**ECG Diagnosis Given:** sinus rhythm. st segments are depressed in i, ii, avl, v5,6. t waves are low or flat in limb leads and v4,5,6. this may be due to lv strain or ischaemia.

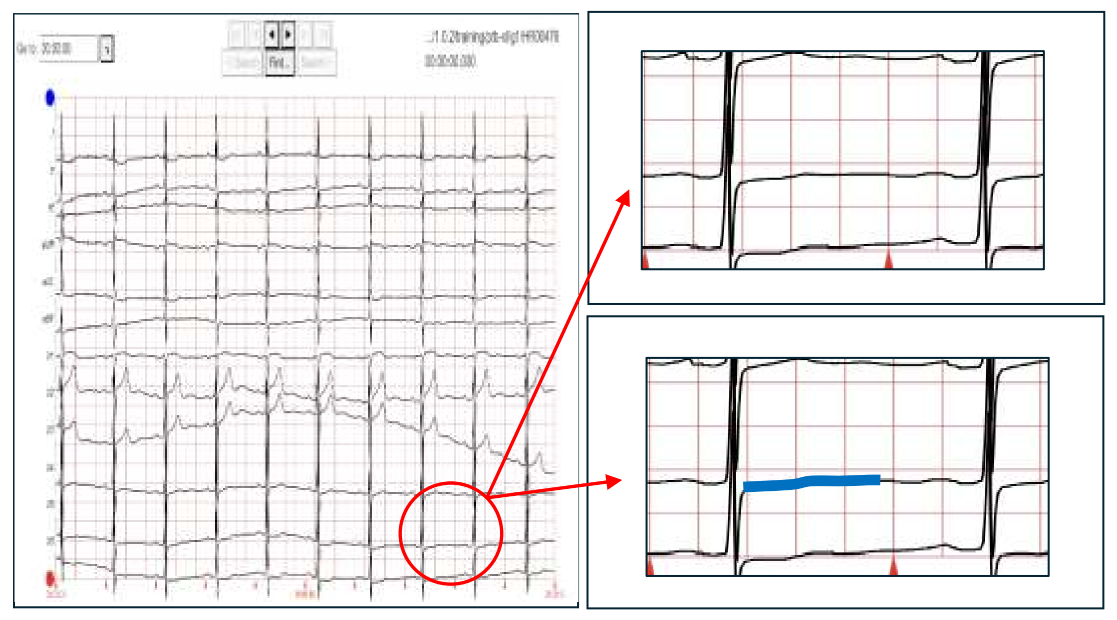

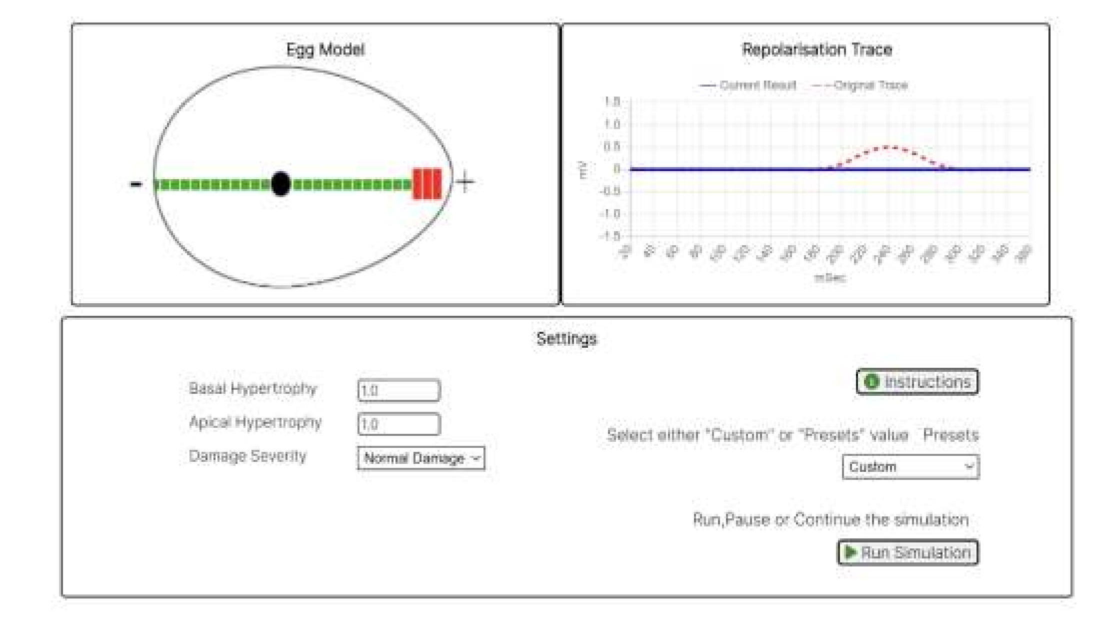

**Commentary**: This is an example of a flat repolarisation trace in lead V5 usually due to pathological causes such as anteroseptal or inferior myocardial infarction/ischaemia, or hypokalaemia.

**Other Similar Example Cases:** training/ptb-xl/g1/HR00069, HR00217, HR00305, HR00495, HR00496, HR00529, HR00535, HR00557, HR00612, HR00692

293

#### Example 4. Inverted T Wave

**ECG ID: training/ptb-xl/g1/HR00271**

**ECG Diagnosis Given:** premature atrial contraction(s). sinus rhythm. left atrial enlargement. qs complexes in v2 and tiny r waves in v3. st segments are depressed and t waves inverted in i, avl, v5, v6 consistent with ischaemic heart disease with old anteroseptal myocardial.

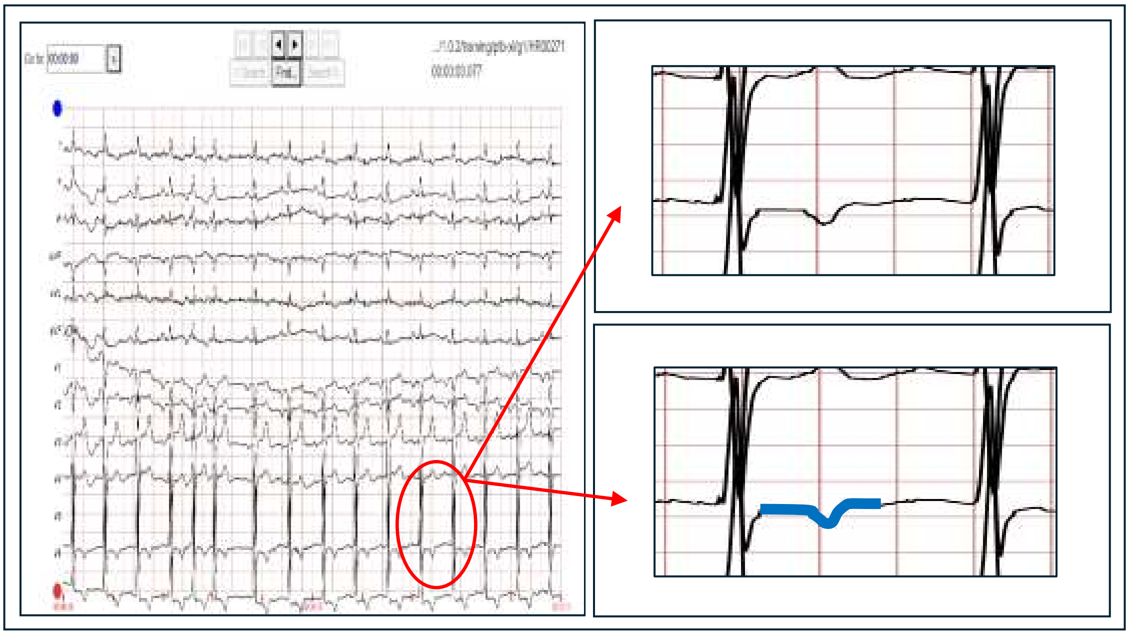

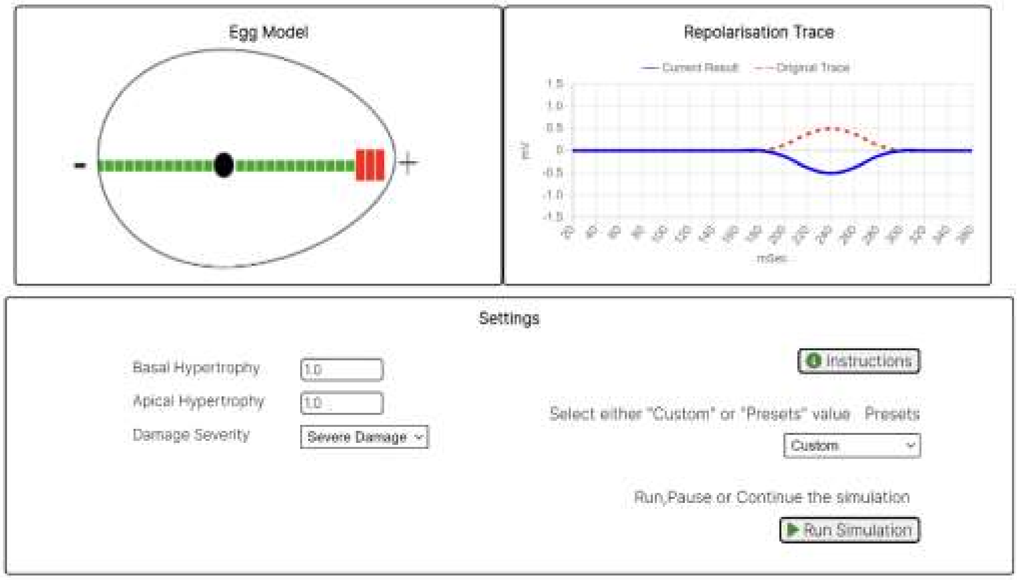

**Commentary**: This is an example of an inverted repolarisation trace in lead V5, usually seen in anteroseptal infarction/ischaemia, bundle branch block, or left ventricular hypertrophy.

**Other Similar Example Cases:** training/ptb-xl/g1/HR00274, HR00281, HR00414, HR00468, HR00564, HR00704, HR00765, HR00914, HR00997

294

#### Example 5. Biphasic T Wave (With Initial Positive Deflection)

**ECG ID: training/ptb-xl/g1/HR00209**

**ECG Diagnosis Given:** sinus rhythm, av block, intraventricular performance disturbance in v1 with steep t wave that changes to negative in v4 v5, pathological ecg.

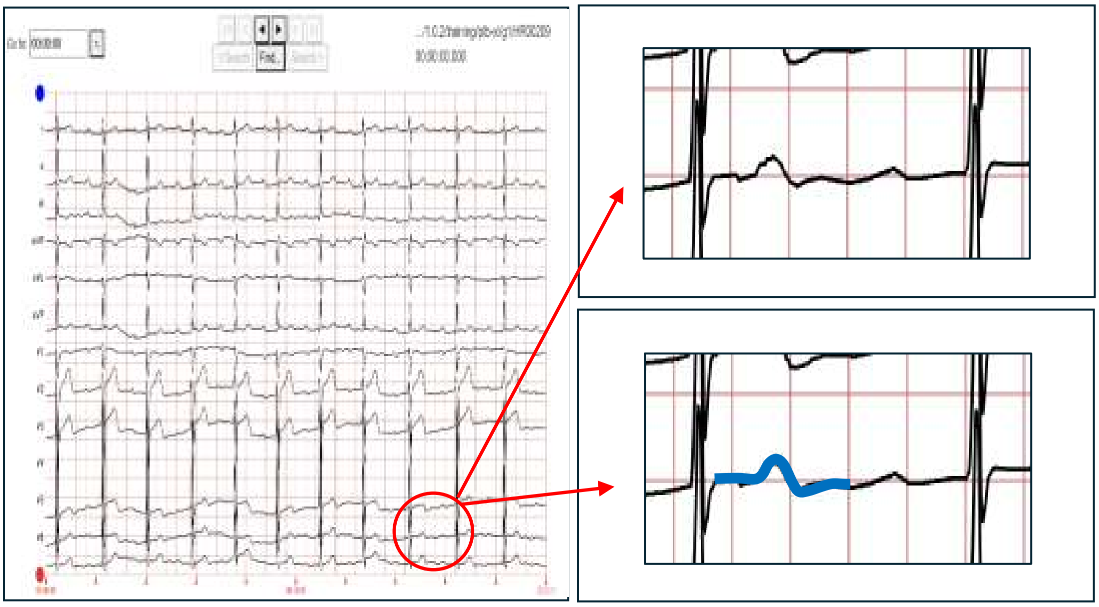

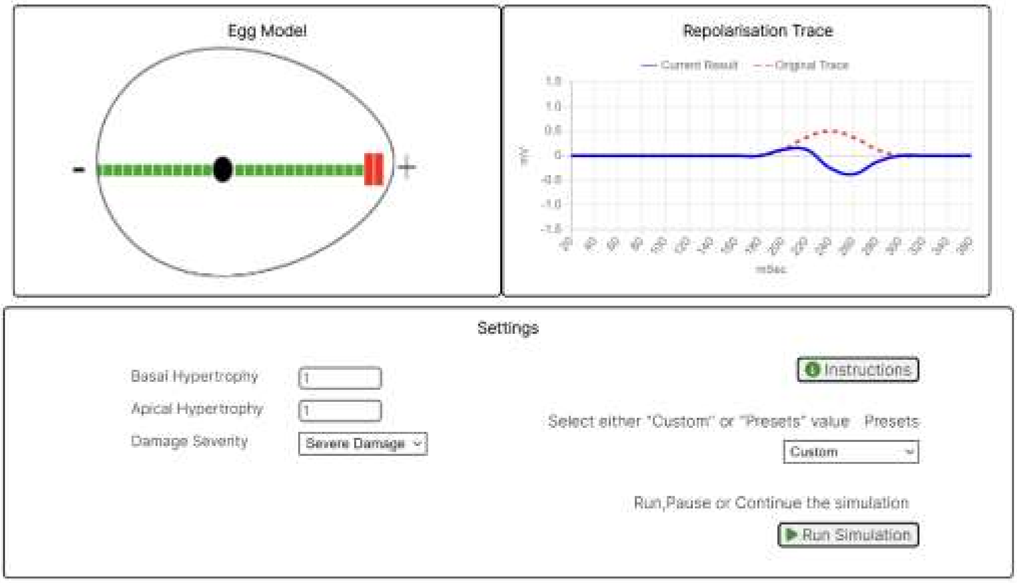

**Commentary**: This is an example of a biphasic repolarisation trace in lead V5 with an initial positive deflection. Usually found in myocardial infarction / ischaemia.

**Other Similar Example Cases:** training/ptb-xl/g1/HR00177, HR00693, HR00898

#### Example 6. Biphasic T Wave (With Initial Negative Deflection)

**ECG ID: training/ptb-xl/g1/HR00519**

**ECG Diagnosis Given:** sinus rhythm extreme left electrical axis left anterior hemiblock st-t lowering, as in inferolateral ischemia or left loading.

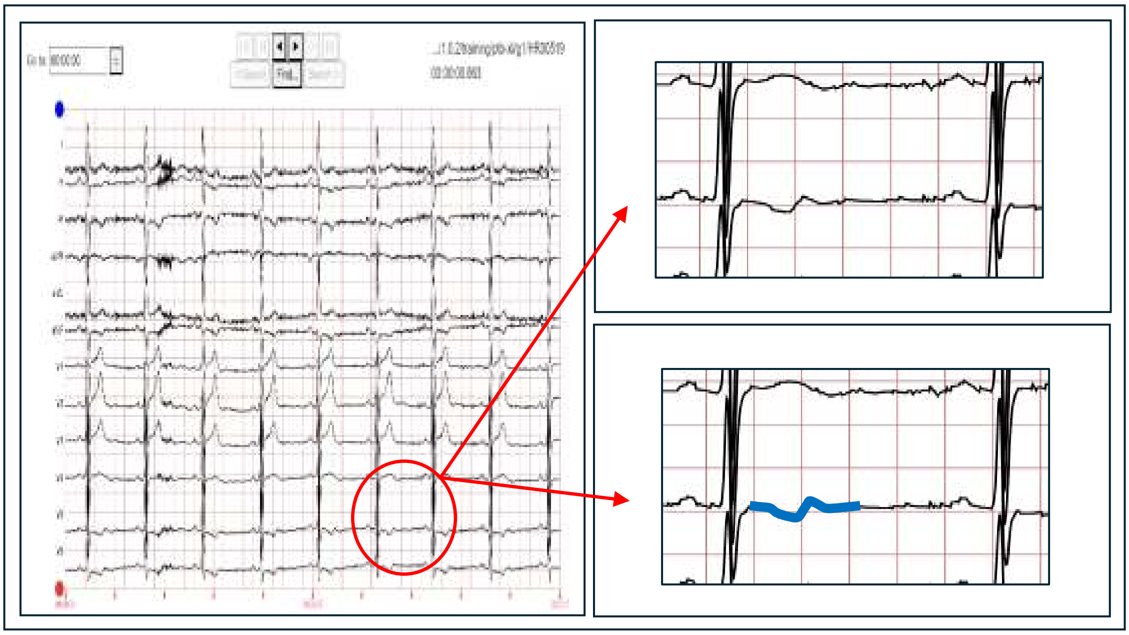

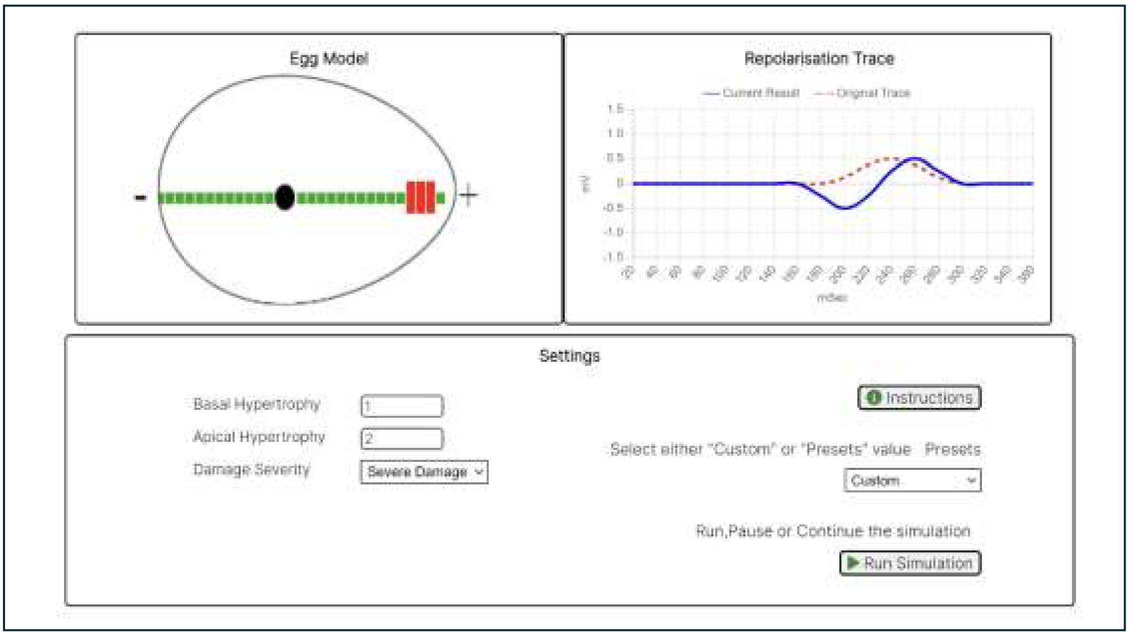

**Commentary**: This is an example of a biphasic repolarisation trace in lead V5 with an initial negative deflection. Usually found in myocardial infarction/ischaemia, or hypokalaemia.

**Other Similar Example Cases:** training/ptb-xl/g1/HR00039, HR00310, HR00530, HR00530, HR00923, HR00941

296

#### Example 7. Bifid T Wave (Camel Hump Shaped)

**ECG ID: training/ptb-xl/g1/HR00512**

**ECG Diagnosis Given:** sinus bradycardia difficult to determine electrical axis, right-sided leg block.

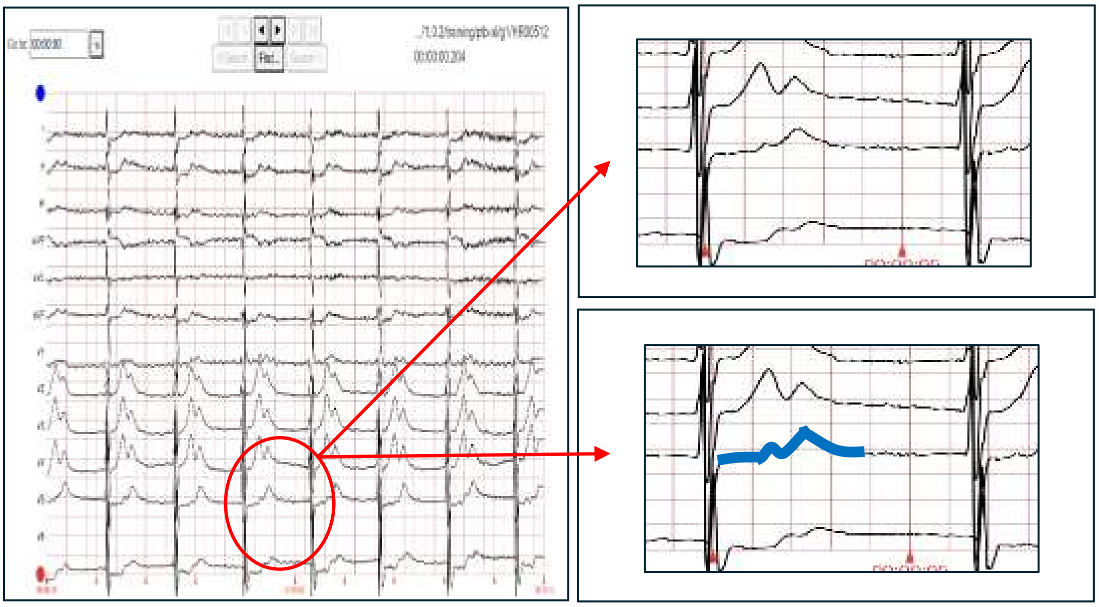

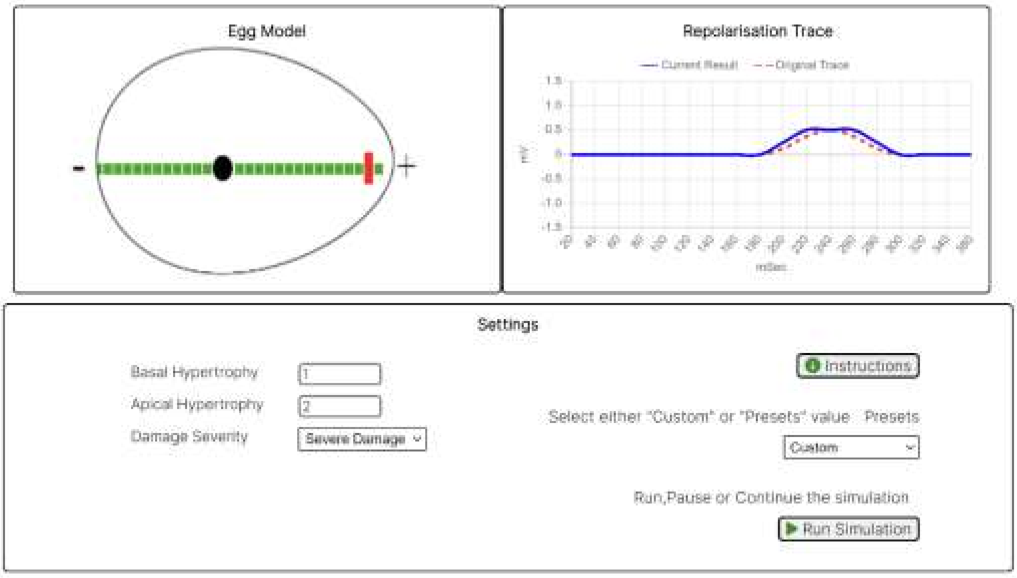

**Commentary**: This is an example of a bifid (camel hump) repolarisation trace in lead V5. They are not common, but can also be seen with fusion of T wave with a prominent U wave or P wave. (The simulated result here can be improved by adding more features as discussed in the conclusion section.)

**Other Similar Example Cases:** training/ptb-xl/g1/HR00458

297

#### 2.3 Part 2. ST Depression and ST Elevation Examples

The Simplified Egg and Changing Yolk model has a symmetrical element and an asymmetrical element as described in the main paper as the 'T zone'.

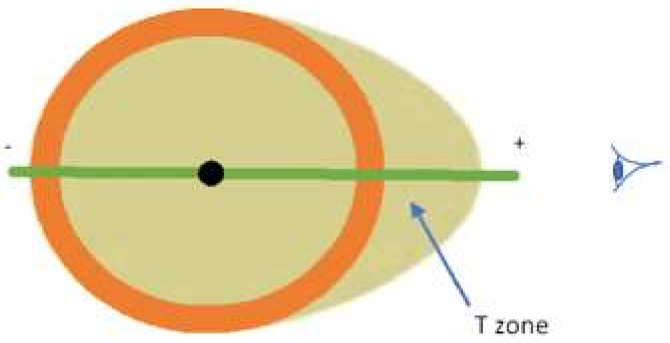

*Figure 1. Representation of the heart in the Simplified Egg and Changing Yolk Model. This figure illustrates a simplified representation of the heart as an egg-shaped 3D object, with the WCT designated at its centre. The green line denotes the vector of lead V5, which, in this instance, passes through the long axis of the egg shape. Signals detected to the right of the WCT for lead V5 register as positive signals, while those on the left side will register as negative signals. The resulting ECG trace generated by the lead is the net result of these positive and negative signals over time. The ST segment of the ventricular repolarisation trace occurs as the signal is expanding out from the WCT over the symmetrical area, whilst the T wave deflection occurs when the repolarisation signal has expanded out into the T zone area.*

The model hypothesizes that the T wave is formed by the asymmetrical part (T zone) which from lead V5 point of view is towards the apex of the heart (egg shape). Since the T wave is 'created' by repolarisation in the T zone, it suggests that T wave shapes can be determined by damage or impaired repolarisation in the heart muscle in the T zone. In this manner we are able to construct the T wave shapes of ‘flat, biphasic and inverted’ T waves. Thus ischaemia or infarct towards the apex of the heart can result in those ‘abnormal’ T wave shapes. Examples 8 to 10 are given to demonstrate the T waves shapes where there is ischaemia (impaired repolarisation) in the lateral part of the heart causing ST depression and altered T wave shapes.

However, the Simplified Egg and Changing Yolk model also suggest that ‘damage’ to the basal part of the heart (distal areas of lead V5 vector), would result in ST elevation (positive deflection), and will not be able to alter the T wave shapes since the T zone is not affected. Hence the model suggest that we would only be able to see ST elevation followed by a positive T wave (see Example 11 below). According to the model it would be difficult to see ST elevation followed by an inverted T wave, or ST elevation followed by biphasic T wave. Indeed in the 1000 ECGs reviewed in this dataset we were unable to find any examples of these.

This is further evidence of the usefulness of the Simplified Egg and Changing Yolk model in predicting how ventricular repolarising traces will appear, as well as a concept for understanding T wave repolarisation shapes.

#### Example 8. ST Depression With A Positive T Wave

**ECG ID: training/ptb-xl/g1/HR00259**

**ECG Diagnosis Given:** sinus rhythm. st segment depression in inferolateral leads suggests ischaemia or left ventricular strain.

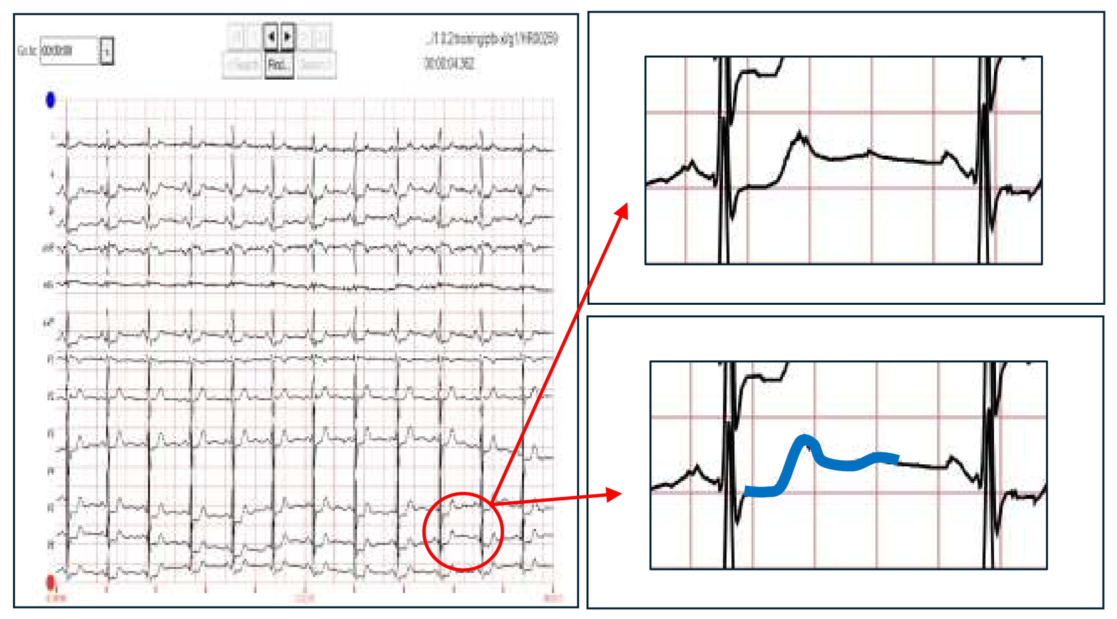

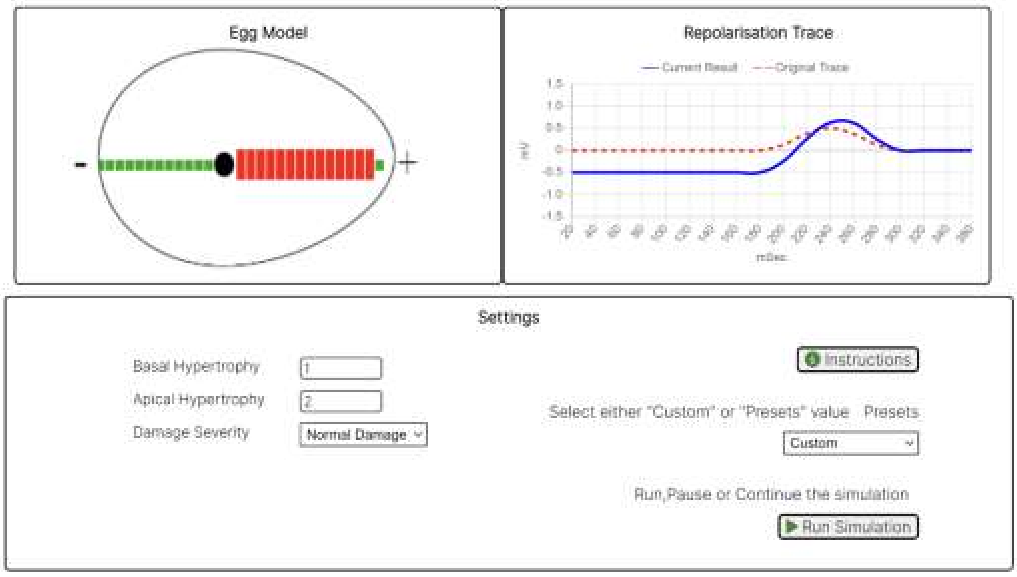

**Commentary**: This is an example of a normal T wave following ST depression in lead V5. These are associated with myocardial lateral ischaemia.

**Other Similar Example Cases:** training/ptb-xl/g1/HR00164, HR00211, HR00223, HR00325, HR00380, HR00425, HR00485, HR00501, HR00513, HR00743

330

#### Example 9. ST Depression With A Biphasic T Wave

**ECG ID: training/ptb-xl/g1/HR00767**

**ECG Diagnosis Given:** sinus rhythm. left atrial enlargement. voltages are high in chest leads suggesting lvh. st segments are depressed and t waves inverted in i, ii, avl, v2-6. this may be due to lv strain or ischaemia.

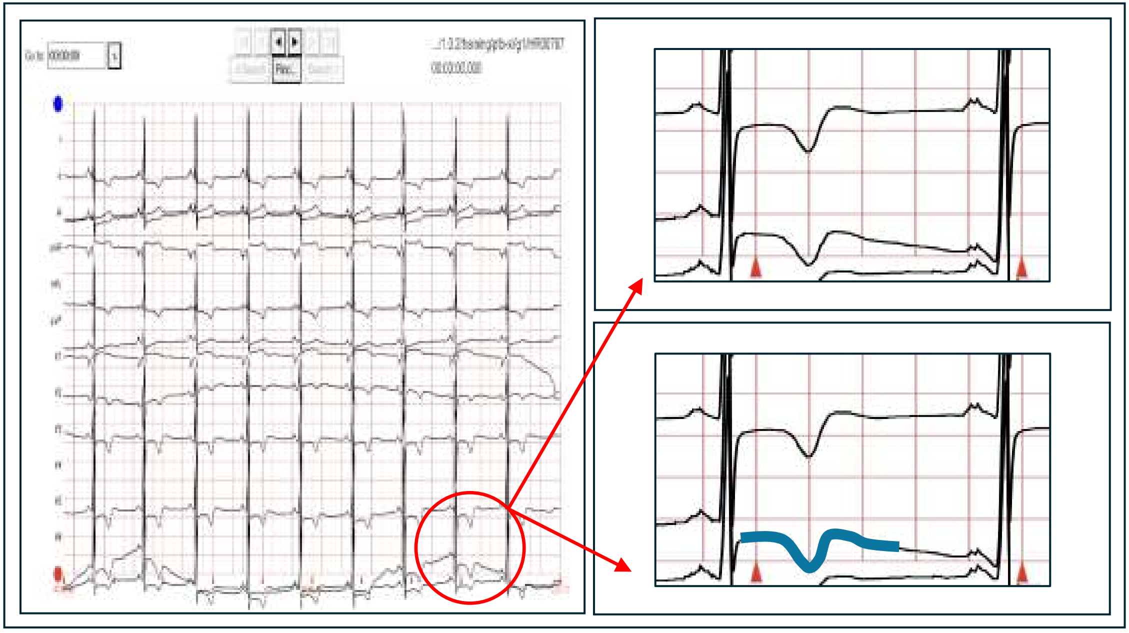

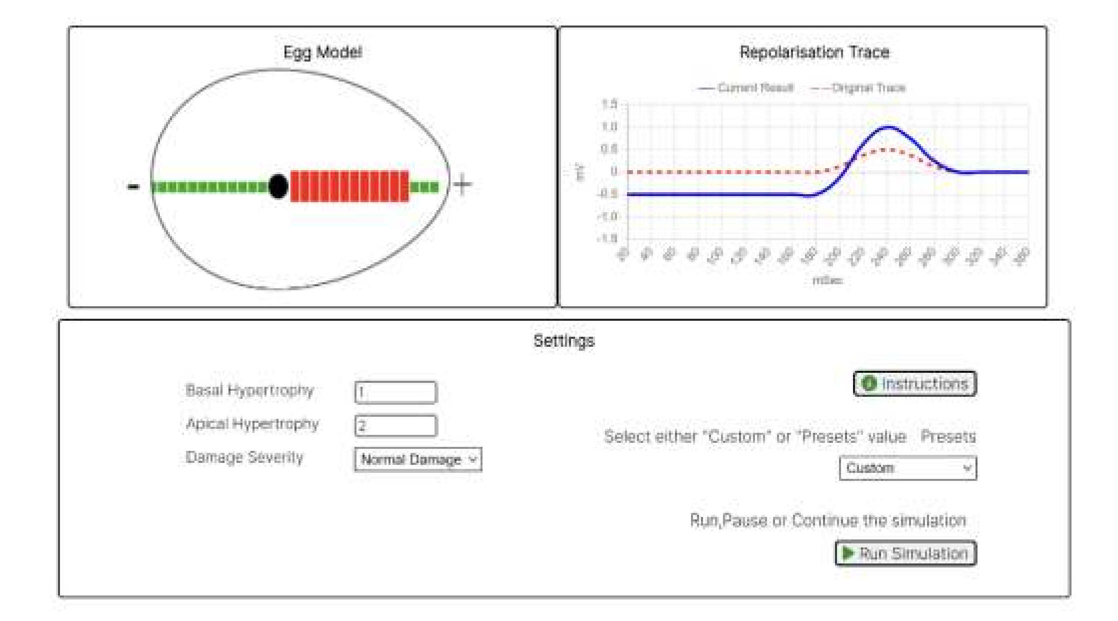

**Commentary**: This is an example of a biphasic T wave following ST depression in lead V5. (The simulated result here can be improved by adding more features to the model as discussed in the conclusion section.)

**Other Similar Example Cases:** training/ptb-xl/g1/HR00282, HR00299, HR00383, HR00423, HR00429, HR00525, HR00545, HR00634, HR00820, HR00934

331

#### Example 10. ST Depression With An Inverted T Wave

**ECG ID: training/ptb-xl/g1/HR00510**

**ECG Diagnosis Given:** sinus rhythm. prolonged pr interval. possible old inferior infarct. sinus bradycardia extreme left electrical axis left anterior hemiblock. st-t decrease, as in anterolateral ischemia or left load t-f–change, as in inferolateral myocardial affection.

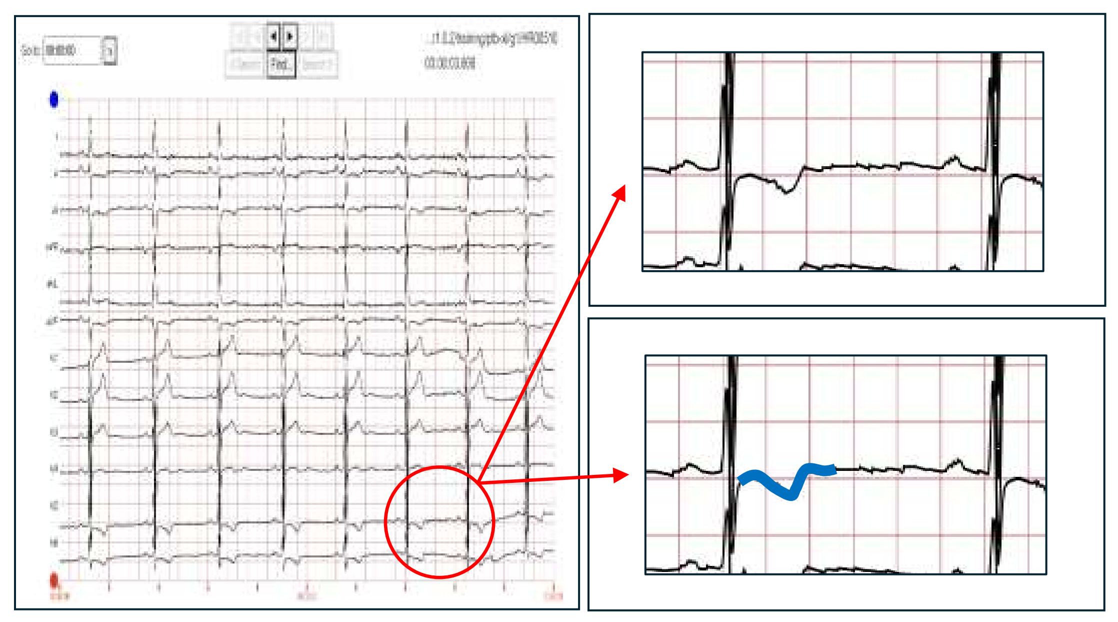

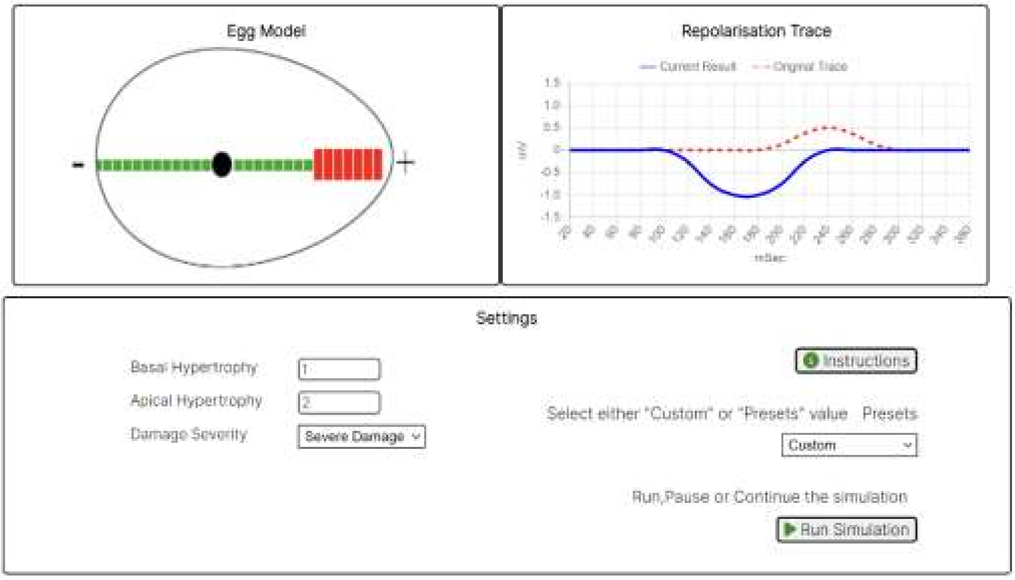

**Commentary**: This is an example of an inverted T wave following ST depression in lead V5. (The simulated result here can be improved by adding more features to the model as discussed in the conclusion section.)

**Other Similar Example Cases:** training/ptb-xl/g1/HR00296, HR00414, HR00461, HR00541, HR00551, HR00585, HR00699, HR00768, HR00828, HR00914

#### Example 11. ST Elevation With A Positive T Wave

**ECG ID: training/ptb-xl/g1/HR00257**

**ECG Diagnosis Given:** premature atrial contraction(s). sinus rhythm. left atrial enlargement. qs complexes in v2. st segments are slightly elevated in v2,3. st segments are depressed in i, avl. t waves are low or flat in i, v5,6 and inverted in avl. consistent with ischaemic heart.

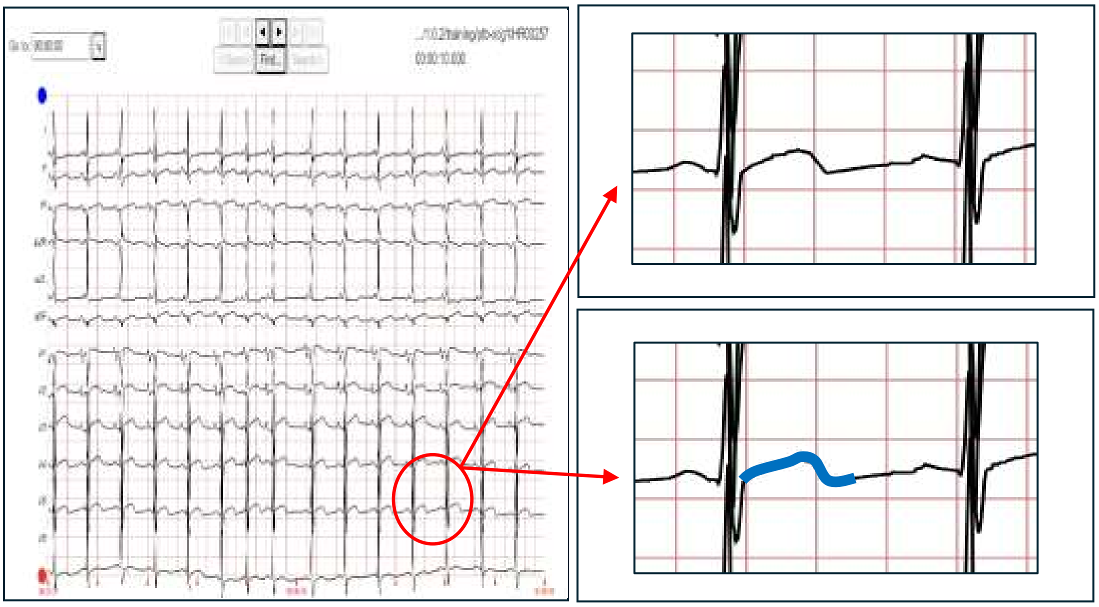

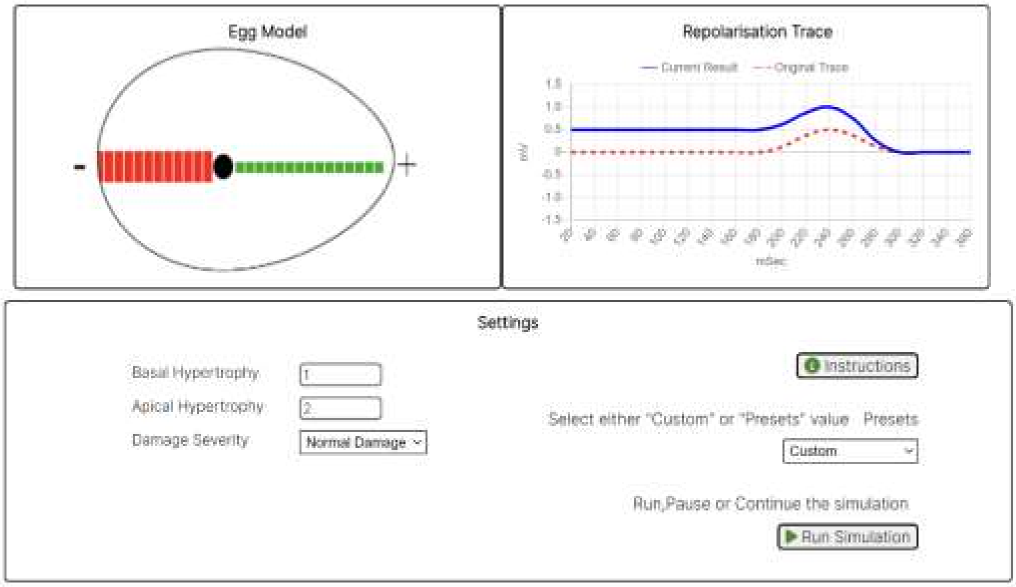

**Commentary**: This is an example of a normal repolarisation trace following ST elevation in lead V5. The model predicts that ST elevation followed by a biphasic or inverted T wave would be difficult to see (unless there is extensive ischaemia / infarction affecting both the basal and lateral aspects of the heart).

**Other Similar Example Cases:** training/ptb-xl/g1/HR00181, HR00184, HR00827, HR00920, HR00947

333

#### 2.4 Part 3. Egg Shape / Heart Orientation Effect on the T Wave

The Simplified Egg and Changing Yolk model has a symmetrical element which defines the ST segment as above, and an asymmetrical element which defines the T wave shape. If however the heart shape is changed so that from lead V5 point of view the egg 'shape' becomes less symmetrical (spherical) with a larger asymmetrical (T zone) element. The effect of this predicted by the Simplified Egg and Changing Yolk model is that the ST segment duration is reduced, and the T wave is seen to start earlier after the QRS complex (See Example 13).

If the heart is rotated around the y axis, then from lead V5 point of view the egg 'shape' becomes more symmetrical with a smaller asymmetrical (T zone) element. If the heart is rotated further, the asymmetrical element disappears and from lead V5 point of view as the heart 'egg' shape becomes spherical. This has the result of producing a flat (or absent) T wave (See Example 14).

Please note that in the Examples 13 and 14 the ECGs are both 'normal', and the different T waves shapes in lead V5 are not from pathological causes.

#### Example 12. Normal T Wave With An Asymmetrical 'Egg' Shape

**ECG ID: training/ptb-xl/g1/HR00014**

**ECG Diagnosis Given:** sinus rhythm. normal ecg.

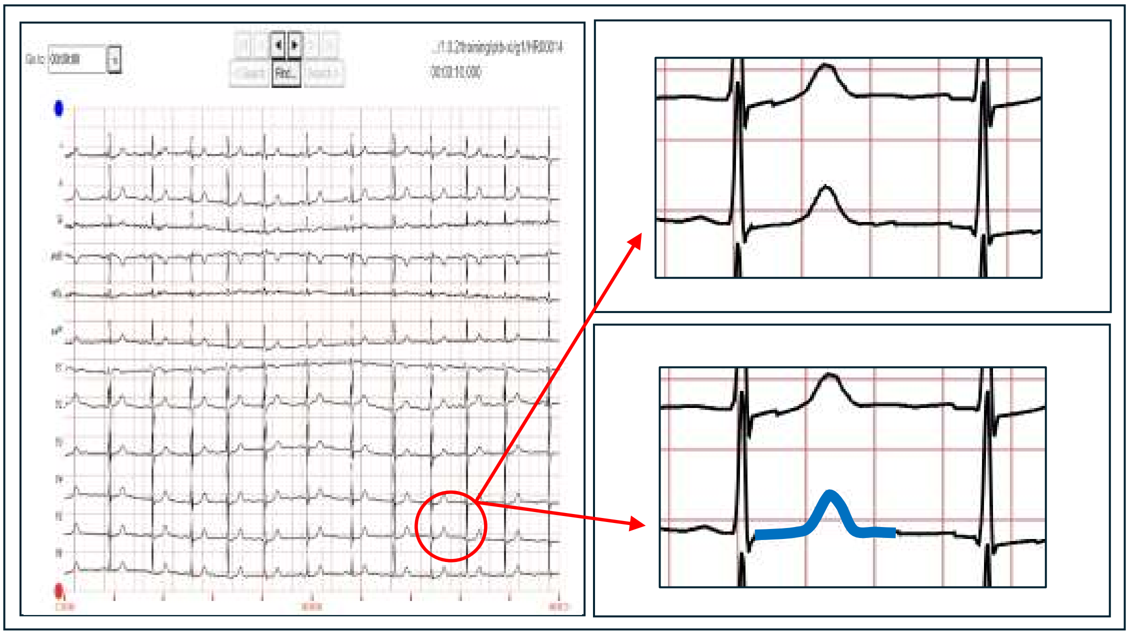

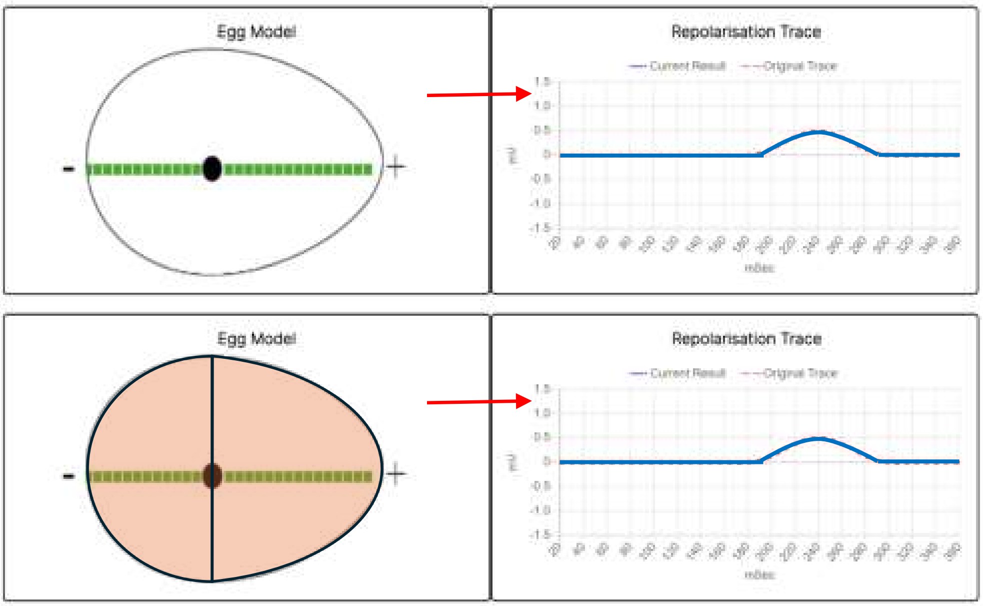

**Commentary**: This is an example of a normal repolarisation trace in lead V5 to demonstrate examples where the T wave deflection starts a short while after the QRS complex. The normal ST segment is flat and represents the 'symmetrical' element of the egg shape. The asymmetrical part generates the T wave.

**Other Similar Example Cases:** training/ptb-xl/g1/HR00005, HR00010, HR00021, HR00037, HR00055, HR00056, HR00066, HR00070, HR00081, HR00082

347

#### Example 13. Normal T Wave With Less Asymmetrical 'Egg' Shape

**ECG ID: training/ptb-xl/g1/HR00009**

**ECG Diagnosis Given:** sinus rhythm. normal ecg.

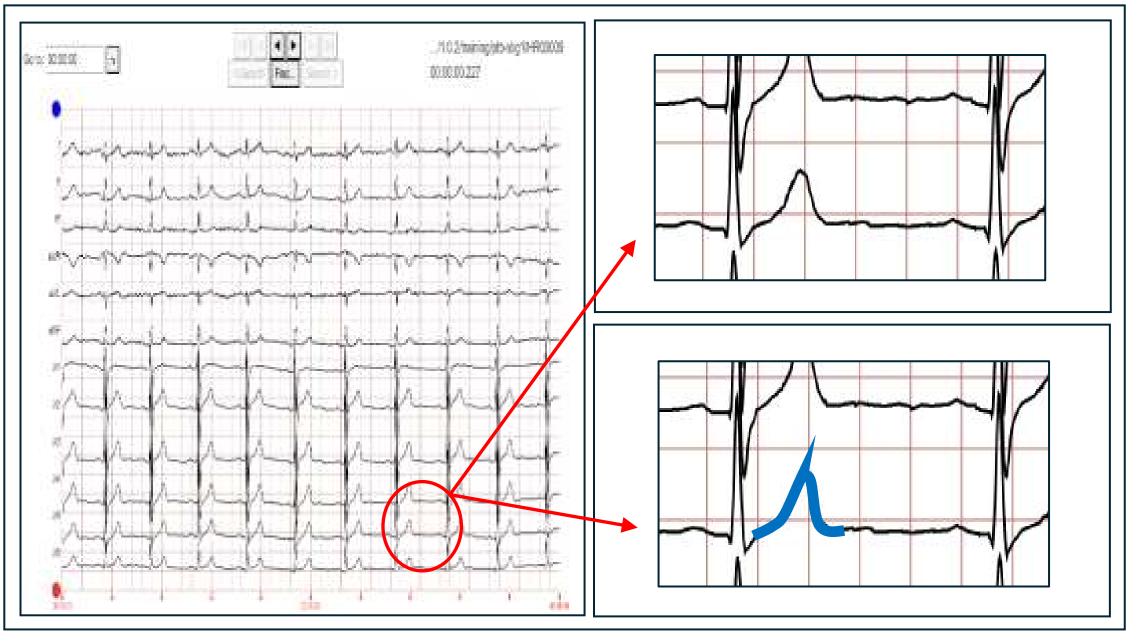

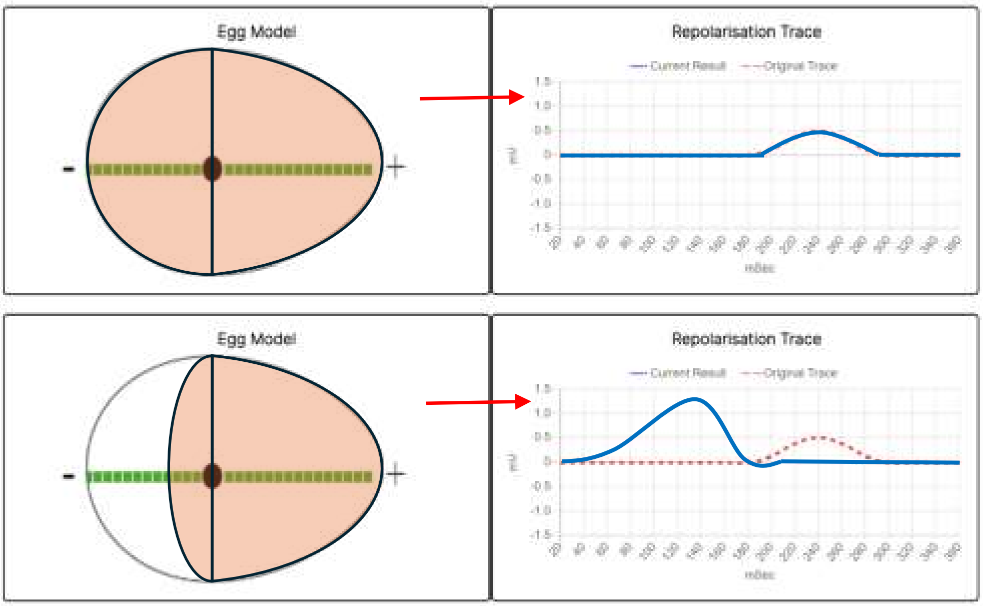

**Commentary**: This is an example of a normal repolarisation trace in lead V5 to demonstrate examples where the T wave deflection starts soon after the QRS complex. The normal ST segment is shortened (or absent) due to the asymmetry.

**Other Similar Example Cases:** training/ptb-xl/g1/HR00002, HR00004, HR00016, HR00035, HR00046, HR00057, HR00075, HR00079, HR00088, HR00095

#### Example 14. Flat T Wave Non-Pathological (Symmetrical ‘Egg’ Shape)

**ECG ID: training/ptb-xl/g1/HR00134**

**ECG Diagnosis Given:** sinus rhythm left electrical axis no definite pathology

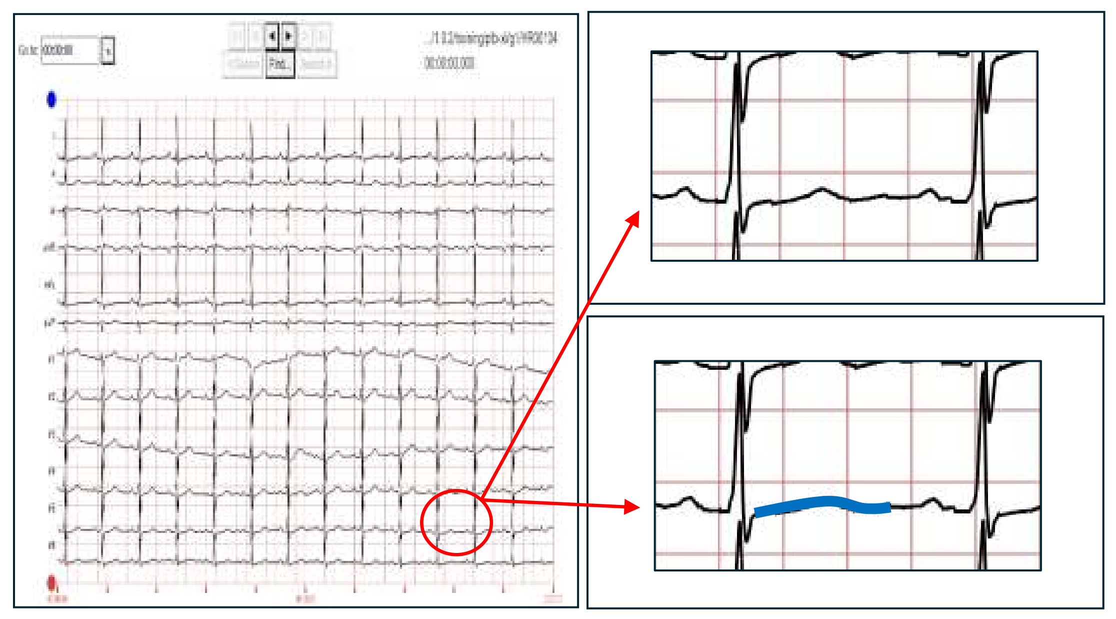

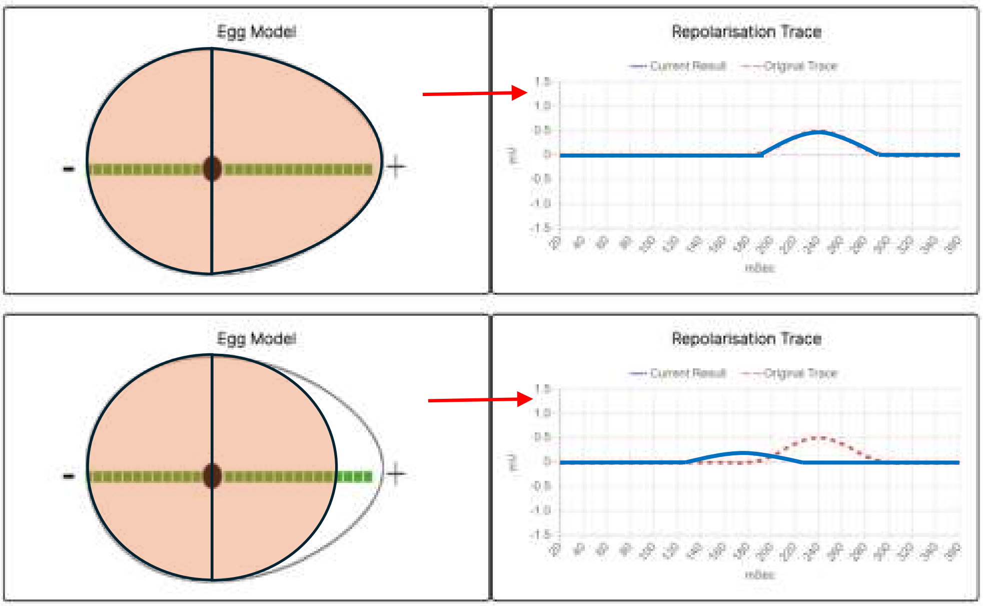

**Commentary**: This is an example of a normal repolarisation trace which has a relatively flat T wave (or absent) in lead V5 due to the orientation of the heart. The resulting shape is symmetrical and so the T wave appears diminished.

**Other Similar Example Cases:** training/ptb-xl/g1/HR00306, HR00682, HR00718, HR00821, HR00974

349

#### 2.5 Conclusion

In this supplementary paper we have demonstrated how the Simplified Egg and Changing Yolk model can be used to describe the normal and pathological T wave patterns that are commonly seen in ECGs. This simple but versatile model is useful in increasing our understanding how the ventricular repolarisation pattern can lead to the various T wave traces on an ECG.

We further proved the usefulness of the Simplified Egg and Changing Yolk model in its ability to predict that inverted and biphasic T wave shapes can be seen in ST depression (impaired repolarisation towards the apex of the heart), but would be difficult to see in ST elevation where the ischaemia is towards the base (right side) of the heart. This is demonstrated by real world the examples from the PTB-XL dataset.

Finally, we have shown how the Simplified Egg and Changing Yolk model can be used to understand how the variations in normal (non-pathological) T waves can be explained. This includes ECGs where the ventricular repolarisation trace in lead V5 has no ST segment, or where the T waves are nearly flat. Although in this model we are looking specifically at lead V5, the principles are applicable to the other ECG leads depending on the ‘shape’ of the heart they are seeing.

Inevitably with a simplified model such as this there are trade-offs to be made in terms of how closely the simulated T waves match the ECG ventricular repolarisation patterns. This we see especially in matching the bifid T wave, and the ST depression followed by a negative or inverted T wave in the simulation. This matching can be improved by increasing the number of blocks and severity of damage levels in this model, however the model presented here is at the ‘simplified’ level in order to explain the principles in an understandable manner.

Having demonstrated the usefulness of this model, in the future we are planning to develop the model further to provide an explanation for the U wave, and to refine the model to include the effects of hypertrophy and coronary artery occlusion in defined territories of the model.

#### References

1. Baumert M, Porta A, Vos MA, et al.: QT interval variability in body surface ECG: measurement, physiological basis, and clinical value: position statement and consensus guidance endorsed by the European Heart Rhythm Association jointly with the ESC Working Group on Cardiac Cellular Electrophysiology. Europace 2016; 18:925–944.

2. Cox AJ, Azeem A, Yeboah J, et al.: Heart Rate–Corrected QT Interval Is an Independent Predictor of All-Cause and Cardiovascular Mortality in Individuals With Type 2 Diabetes: The Diabetes Heart Study. Diabetes Care 2014; 37:1454–1461.

3. Portaluppi F, Hermida RC: Circadian rhythms in cardiac arrhythmias and opportunities for their chronotherapy. Advanced Drug Delivery Reviews 2007; 59:940–951.

4. Sztajzel J: Heart rate variability: a noninvasive electrocardiographic method to measure the autonomic nervous system. Swiss Med Wkly [Internet] 2004 [cited 2024 Dec 11]; . Available from: https://smw.ch/index.php/smw/article/view/411

5. Chiladakis J, Kalogeropoulos A, Arvanitis P, Koutsogiannis N, Zagli F, Alexopoulos D: Heart Rate-Dependence of QTc Intervals Assessed by Different Correction Methods in Patients with Normal or Prolonged Repolarization: HEART RATE-DEPENDENCE OF QTc INTERVALS. Pacing and Clinical Electrophysiology 2009; 33:553–560.

6. Rautaharju PM, Surawicz B, Gettes LS: AHA/ACCF/HRS Recommendations for the Standardization and Interpretation of the Electrocardiogram. Journal of the American College of Cardiology 2009; 53:982–991.

7. Antzelevitch C: Transmural dispersion of repolarization and the T wave. Cardiovascular Research 2001; 50:426–431.

8. Yan G-X, Antzelevitch C: Cellular Basis for the Normal T Wave and the Electrocardiographic Manifestations of the Long-QT Syndrome. Circulation 1998; 98:1928–1936.

9. Postema P, Wilde A: The Measurement of the QT Interval. CCR 2014; 10:287–294.

10. Drew BJ, Califf RM, Funk M, et al.: Practice Standards for Electrocardiographic Monitoring in Hospital Settings: An American Heart Association Scientific Statement From the Councils on Cardiovascular Nursing, Clinical Cardiology, and Cardiovascular Disease in the Young: Endorsed by the International Society of Computerized Electrocardiology and the American Association of Critical-Care Nurses. Circulation 2004; 110:2721–2746.

11. Goldenberg I, Moss AJ, Zareba W: QT Interval: How to Measure It and What Is "Normal." Cardiovasc electrophysiol 2006; 17:333–336.

12. De Oliveira Neto NR, De Oliveira WS, Campos Pinto GD, De Oliveira ESR, Da Silveira Barros M das ND: A Practical Method for QTc Interval Measurement. Cureus 2020; 12:e12122.

13. Vandenberk B, Vandael E, Robyns T, et al.: Which QT Correction Formulae to Use for QT Monitoring? JAHA 2016; 5:e003264.

14. Al-Khatib SM, LaPointe NMA, Kramer JM, Califf RM: What Clinicians Should Know About the QT Interval. JAMA [Internet] 2003 [cited 2024 Dec 10]; 289. Available from: http://jama.jamanetwork.com/article.aspx?doi=10.1001/jama.289.16.2120

15. El-Sherif N, Turitto G: Electrolyte disorders and arrhythmogenesis. Cardiol J 2011; 18:233–245.

16. Whang R, Whang DD, Ryan MP: Refractory Potassium Repletion: A Consequence of Magnesium Deficiency. Arch Intern Med 1992; 152:40.

17. Pickham D, Helfenbein E, Shinn JA, et al.: High prevalence of corrected QT interval prolongation in acutely ill patients is associated with mortality: Results of the QT in Practice (QTIP) Study*. Critical Care Medicine 2012; 40:394–399.

18. Bernardi M, Calandra S, Colantoni A, et al.: Q-T interval prolongation in cirrhosis: Prevalence, relationship with severity, and etiology of the disease and possible pathogenetic factors. Hepatology 1998; 27:28–34.

19. Ziegler D: Diabetic cardiovascular autonomic neuropathy: Prognosis, diagnosis and treatment. Diabetes Metab Rev 1994; 10:339–383.

20. Perez Alday EA, Gu A, Shah A, et al.: Classification of 12-lead ECGs: The PhysioNet/Computing in Cardiology Challenge 2020 [Internet]. PhysioNet, [cited 2024 Dec 11],. Available from: https://physionet.org/content/challenge-2020/1.0.2/

### Simplified Egg and Changing Yolk Model
### User Manual (version 1.0)

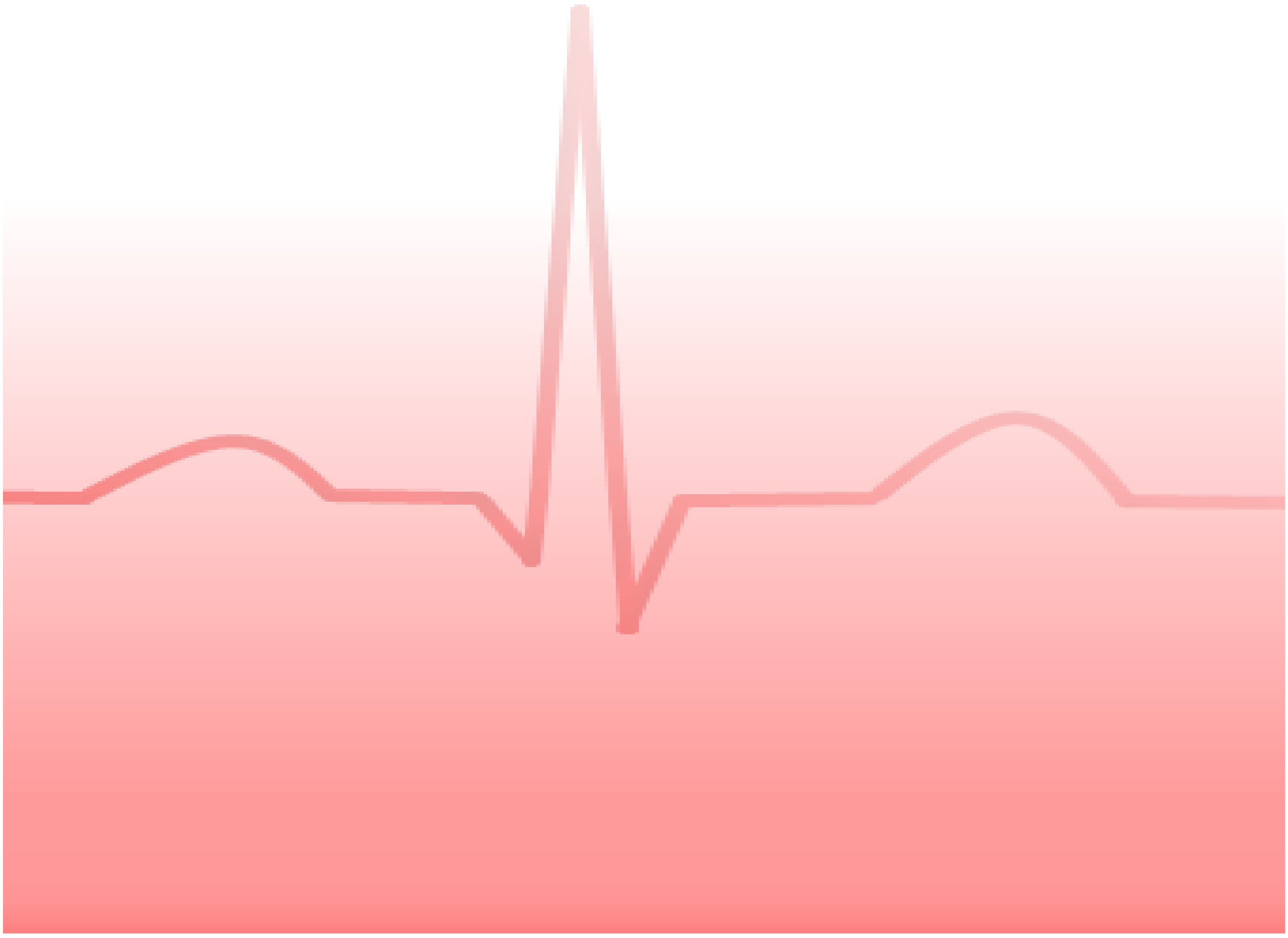

**Contents** **Page No.**

#### 1. Overview

This web tool models the ventricular repolarisation, which is currently thought to be the ST and T wave segments on an electrocardiograph (ECG) trace. In the model, the heart is simplified to a mass of muscle in the form of an 'egg' shape. Here, the vector for lead V5 is seen as passing through the long axis of the egg shape. The repolarisation trace represents the calculated electrical deflection as seen in lead V5 over time. The model allows you to allocate the severity and location of any hypertrophy and/or damage to see the effects on the repolarisation trace.

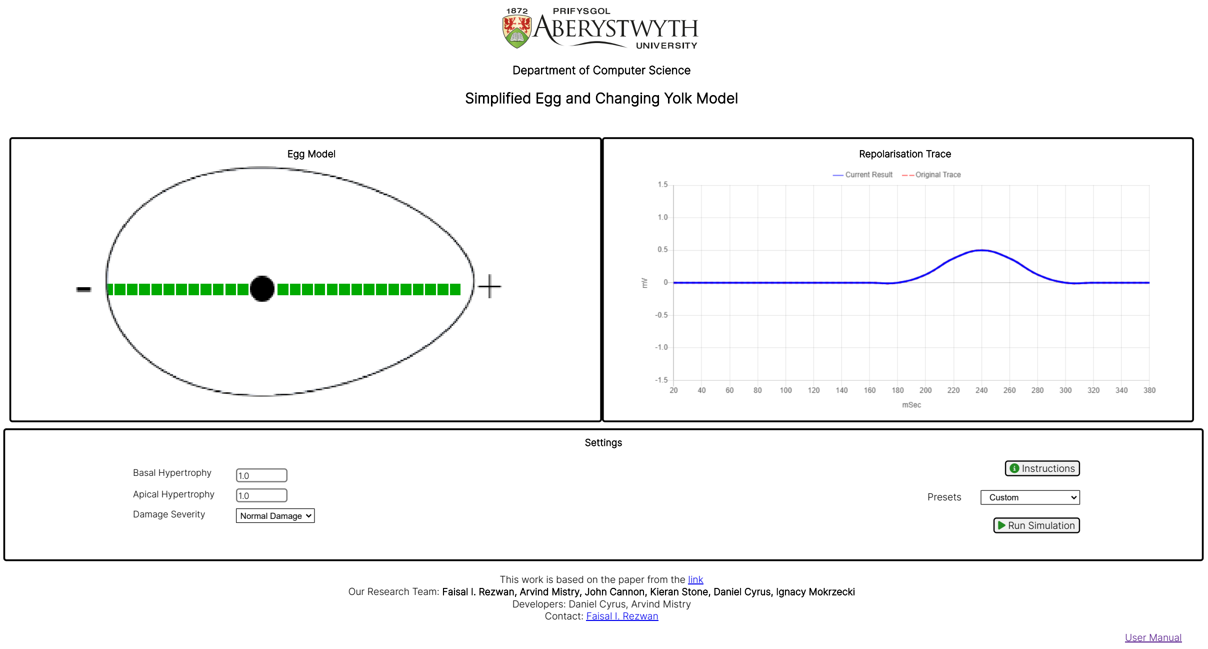

#### 2. Instructions

In this web tool, you can select the 'Instructions' button at any time to see the different elements and functions of the model. These instructions will appear in green pop-up boxes. The pop-up boxes disappear when any other selection is subsequently made.

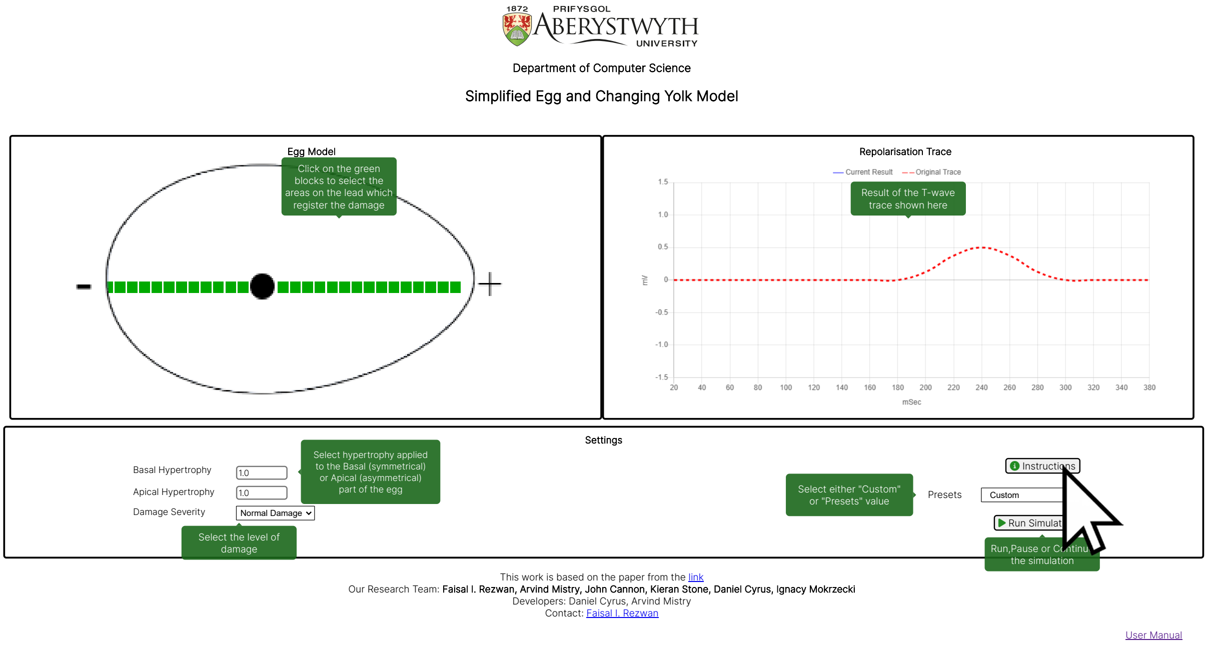

#### 3. Damage Location Selection

You can allocate the areas where any damage is registered on lead V5 by clicking the selected green blocks on the egg. The green block will change to red indicating where the damage is registered. By clicking the (red) block again, you can undo any damage and the block will reset to green.

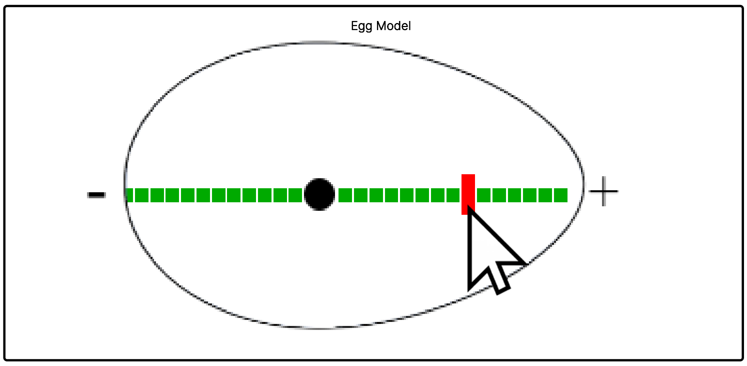

#### 4. Choosing Damage Severity

You can select either 'Normal' or 'Severe' levels of damage to be applied to the damaged areas of egg. This allows this model to allocate a higher level of damage for demonstrating certain conditions, such as flattened and inverted T waves.

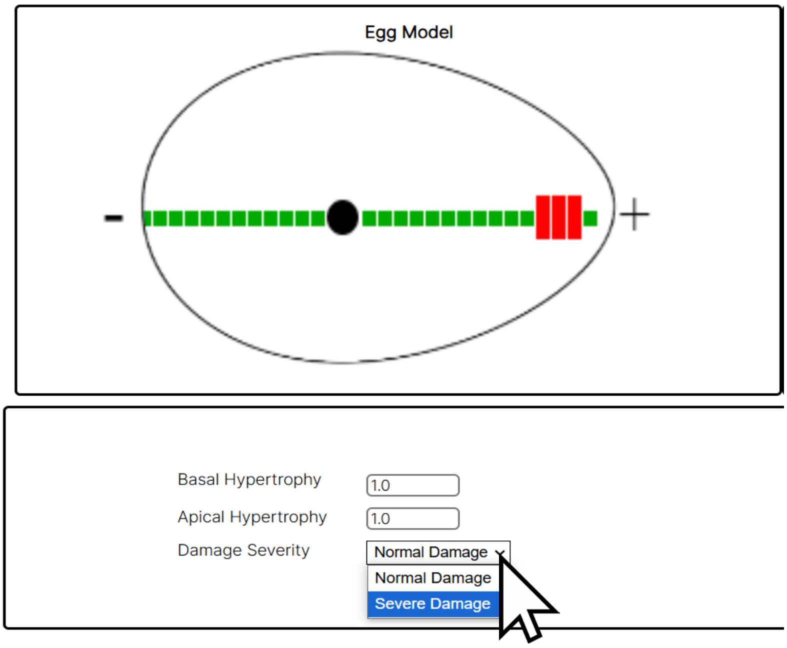

#### 5. Hypertrophy Allocation

You can select the level of 'Basal' and or 'Apical' hypertrophy to be applied to the model. Basal applies hypertrophy to the 'symmetrical' part or body of the egg shape. Apical applies hypertrophy to the 'asymmetrical' to apex part of the egg shape.

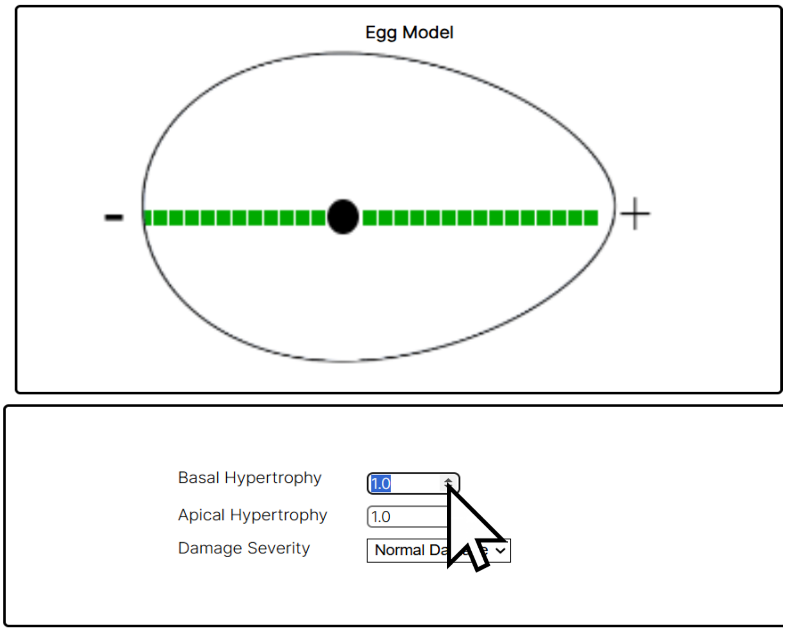

The range of hypertrophy that can be allocated in this simplified model is between 1.0 and 2.0. Allocating 1.0 for either Basal or Apical Hypertrophy means that those parts are normal and there is no allocated hypertrophy at those sites. By allocating more than 1.0 hypertrophy means that area of the model has some hypertrophy.

For example, if hypertrophy is applied to the apical part of the egg model **without** any allocated damage, then this will result in a higher T wave in the repolarisation trace. If any damage is allocated to a hypertrophied part of the egg, then this will have a greater 'damage' effect on the repolarisation trace.

#### 6. Presets Selection

As an alternative to 'Custom' adjusting the settings, you can pre-select damaged and hypertrophy areas by selecting one of eleven 'Presets'.

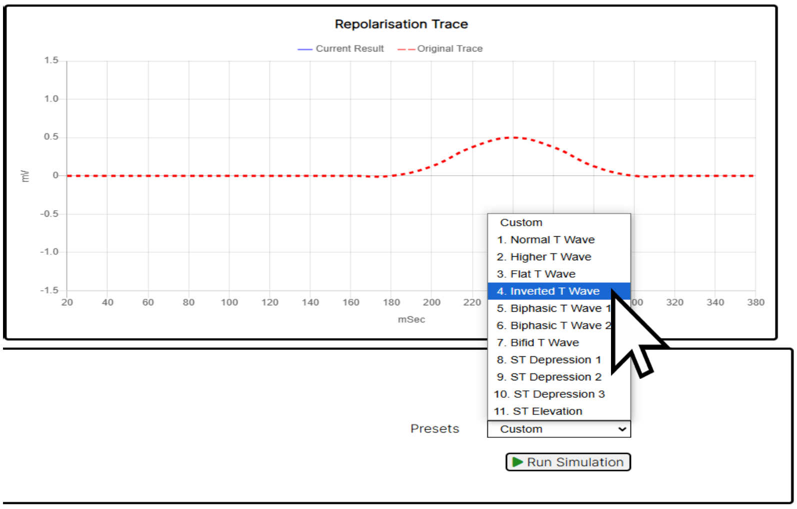

The theory underpinning these Presets are discussed in the paper linked to this model.

#### 7. Running the Simulation

When the 'Run Simulation' button is selected the model will calculate and display the resulting repolarisation trace for this simplified model. Deviations from the original repolarisation trace indicate hypertrophy and/or damage to the heart.

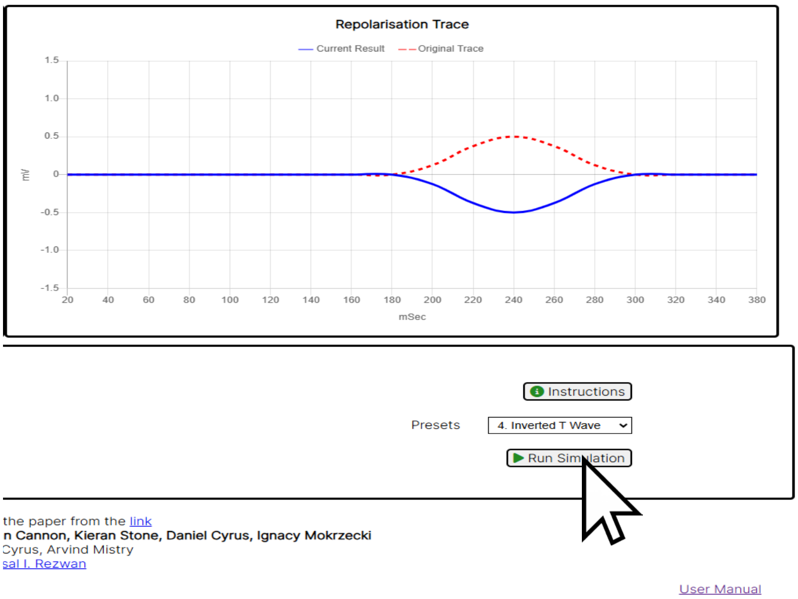

#### 8. Hot Tips for Using the Model

When selecting the 'Damage' blocks, it is best to select one, two or three adjacent blocks on one side of the egg to mimic a localised area of damage.

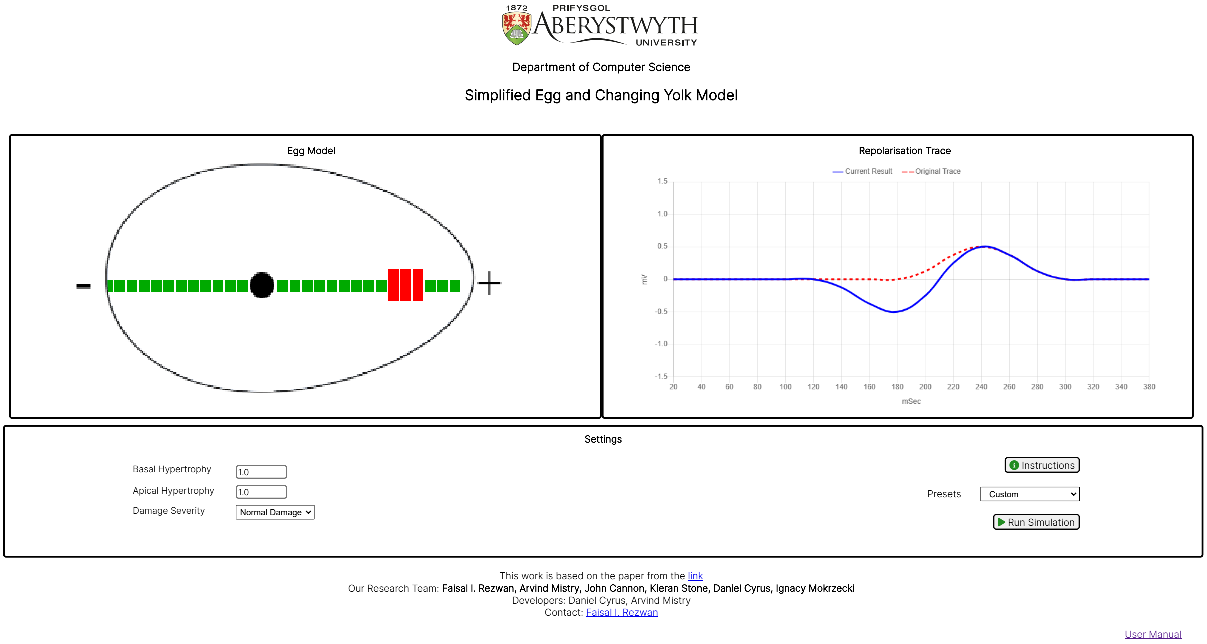

It is possible to have 'Damage' blocks on both sides of the egg shape, which results in various specialised configuration of the repolarisation trace. However, explanations of circumstances, when this would occur, are outside of the remit of this web tool. Please contact the research team for more detailed explanation.

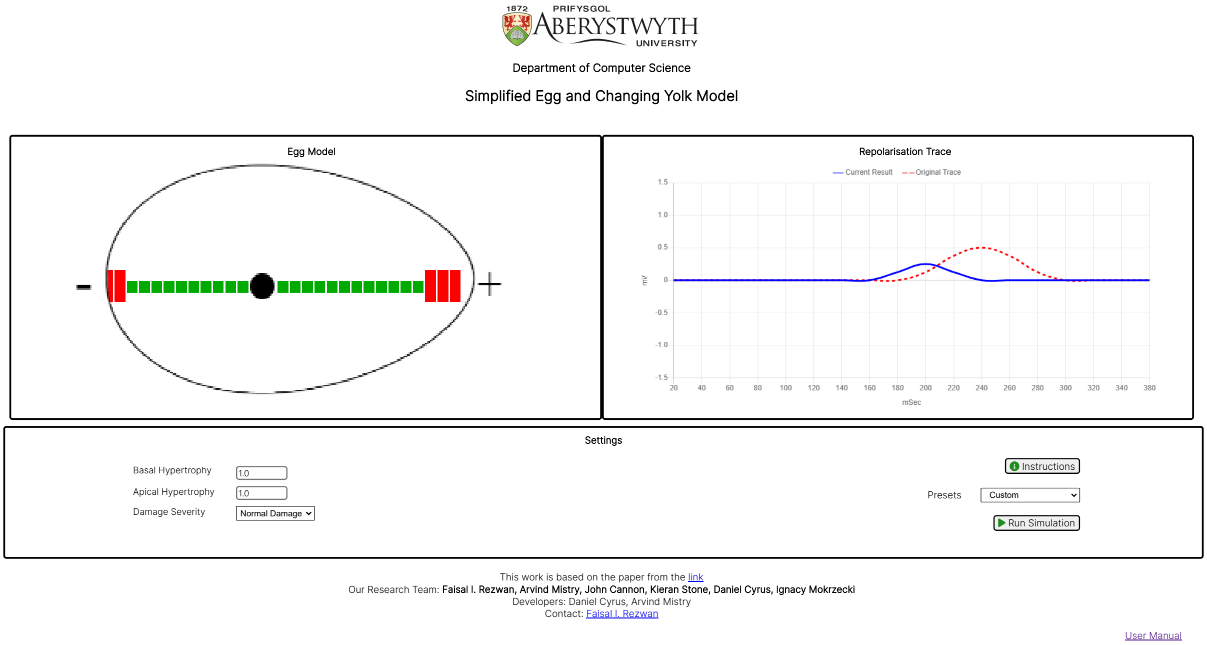

#### 9. Known issue(s):

The texts inside green instruction pop-ups do not load properly in safari web browser in iOS. They only appear after refreshing the web tool.

#### Research Team and Contact Details

This "Simplified Egg and Changing Yolk Model" has been developed by Aberystwyth University Computer Science Department as a part of the 3D ECG Project.

Developers: Daniel Cyrus and Arvind Mistry

Research Team: Faisal I. Rezwan (Project Lead), Arvind Mistry, John Cannon, Kieran Stone, Daniel Cyrus and Ignacy Mokrzecki

For any queries or support, please contact:
